# Autoantibodies against nephrin elucidate a novel autoimmune phenomenon in proteinuric kidney disease

**DOI:** 10.1101/2021.02.26.21251569

**Authors:** Andrew J.B. Watts, Keith H. Keller, Gabriel Lerner, Ivy Rosales, A. Bernard Collins, Miroslav Sekulic, Sushrut S. Waikar, Anil Chandraker, Leonardo V. Riella, Mariam P. Alexander, Jonathan P. Troost, Junbo Chen, Damian Fermin, Jennifer Lai Yee, Matthew Sampson, Laurence H. Beck, Joel M. Henderson, Anna Greka, Helmut G. Rennke, Astrid Weins

**Affiliations:** Department of Pathology, Brigham and Women’s Hospital, and Harvard Medical School, Boston, MA; Renal Division, Department of Medicine, Brigham and Women’s Hospital, and Harvard Medical School, Boston, MA; Department of Pathology, Boston Medical Center and Boston University, Boston, MA; Section of Nephrology, Department of Medicine, Boston Medical Center and Boston University, Boston, MA; Department of Pathology, Massachusetts General Hospital, and Harvard Medical School, Boston, MA; Department of Pathology and Cell Biology, Columbia University College of Physicians and Surgeons; Department of Laboratory Medicine and Pathology, Mayo Clinic, Rochester, MN; Division of Nephrology, Department of Pediatrics, University of Michigan, Ann Arbor, MI; Division of Medicine, Department of Pediatrics, University of Michigan School of Medicine, Ann Arbor, MI; Department of Medicine/Pediatric Nephrology, Boston Children’s Hospital, and Harvard Medical School, Boston, MA; Broad Institute of MIT and Harvard, Cambridge, MA

## Abstract

Dysfunction of podocytes, cells critical for glomerular filtration, underlies proteinuria and kidney failure. Genetic forms of proteinuric kidney disease can be caused by mutations in several podocyte genes, including nephrin, a critical component of the kidney filter. In contrast, the etiology of acquired acute-onset nephrotic syndrome has remained elusive. Here we identify autoantibodies against nephrin in serum and glomeruli of a subset of adults and children with non-congenital acute nephrotic syndrome. Our findings align with published experimental animal studies and elucidate a novel autoimmune phenomenon in proteinuric kidney disease interfering with glomerular filter integrity.

## Introduction

Diffuse podocytopathy with minimal changes (Minimal Change Nephrotic Syndrome, MCNS) is an important and common pathologic diagnosis in adults and children with nephrotic syndrome (NS). It is characterized by minimal changes by light microscopy, but extensive injury to glomerular podocytes with diffuse foot process effacement (FPE) and loss of slit diaphragms (SD) by electron microscopy (EM) in the absence of electron dense deposits^1^. The consequence of these alterations is massive proteinuria secondary to failure of the glomerular filtration barrier (GFB), whose integrity is critically dependent on the specialized junctional SD protein complex linking the interdigitating podocyte foot processes.

Nephrin is an essential component of the SD^2,3^, as illustrated by genetic mutations in nephrin, that cause complete lack of cell surface localization, underlying Congenital Nephrotic Syndrome of the Finnish Type (CNF)^4,5^. In contrast to congenital NS with an established genetic basis, the cause of non-congenital NS in both children and adults remains largely unknown. There is strong evidence supporting immune dysregulation with a potential causative circulating factor, however its identity has remained elusive^6,7^. Glucocorticoids are effective at inducing remission, however relapse, steroid dependence and intolerance are common, often requiring alternative immunosuppressive agents^8^. In those patients with steroid dependent NS who progress to end stage kidney disease (ESKD) and require kidney transplantation, the disease can promptly recur in the allograft^1^.

The recent discovery that anti-CD20 B-cell targeted therapies are effective in children with frequently relapsing or steroid-dependent NS^9-11^ and in adults^12^ suggests a potential autoantibody-mediated etiology. However, this possibility is hard to reconcile with the traditional view of MCNS lacking IgG deposition on renal biopsy^13^. Whilst diffuse podocyte-associated IgG is described in MCD, it is minimal compared to that seen in membranous nephropathy (MN), and in the absence of deposits by EM is generally attributed to non-specific protein resorption of little significance^14^.

Antibodies targeting this essential SD component nephrin have been shown to cause massive proteinuria when administered in animal models^15-17^ and when they arise as alloantibodies following kidney transplantation in children with CNF and complete nephrin deficiency^18^. In both animal models^15,16^ and cultured podocytes^7,19^, anti-nephrin antibodies cause a redistribution of nephrin that is identical to that observed in renal biopsies of patients with NS^20,21^. This redistribution of nephrin away from the SD has been proposed as a mechanism to explain the proteinuria in these patients; however, the cause of this redistribution remains unknown.

Differing from classic immune complex deposition diseases such as membranous nephropathy, the phenomenon of functional autoantibody-mediated disruption of a junctional adhesion complex is well described for the blistering skin condition pemphigus^22^. In this disease, autoantibodies directly bind desmogleins (dsg), a critical constituent of the desmosomal cell adhesion complex, analogous to nephrin in the SD complex, and cause redistribution of dsg away from the cell surface with consequent loss of desmosomal integrity and ultimately cell-cell interactions^22^.

Taken together these observations led us to hypothesize that autoantibodies against nephrin may underlie non-congenital MCNS by affecting the integrity of the SD complex.

## Results

To first determine whether circulating autoantibodies against nephrin are detectable in the serum of patients with biopsy proven MCD and no known genetic basis (lacking known pathogenic variants in established Mendelian NS genes), we evaluated serum obtained from the Nephrotic Syndrome Study Network (NEPTUNE) longitudinal cohort study^23^ consisting of 41 (66%) children and 21 (34%) adults (Table S1). We developed an indirect enzyme-linked immunosorbent assay (ELISA) using a recombinant, affinity purified extracellular domain of human nephrin (hNephrin_G1059_) (Fig. S1) and established a threshold for anti-nephrin antibody (ab) positivity, based on the maximum titer in a healthy control population (n=30) (Fig. 1a). Evaluation of the earliest serum sample obtained during active disease (urine protein to creatinine ratio, UPCR > 3g/g) revealed that 18 (29%) of 62 patients, with an equal number of adults and children, were positive for autoantibodies against nephrin (Fig. 1a). Control sera from 53 (98%) of 54 patients who tested positive for anti-hPLA_2_R antibodies, by clinically validated ELISA and indirect immunofluorescence test (IIFT) assays, were negative for anti-nephrin ab (Fig. 1a).

**Figure 1:**
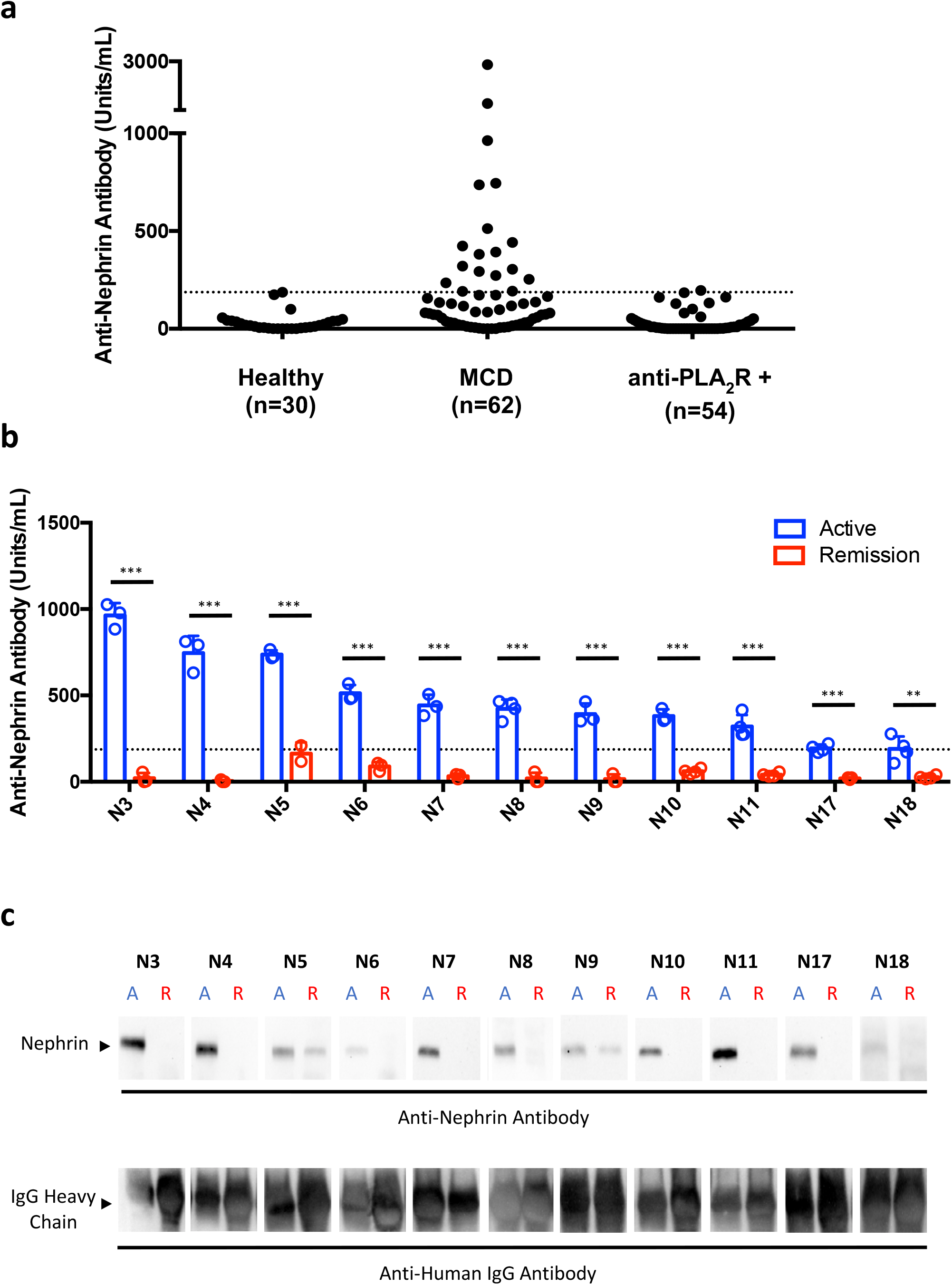
Circulating autoantibodies against nephrin are present in a subset of MCD patients from the NEPTUNE study cohort and correlate with disease activity. **(a)** Antibodies against the extracellular domain of recombinant human nephrin (hNephrin_G1059_) were measured by indirect ELISA. Antigen specific binding was determined by subtracting the average OD_450nm_ of duplicate uncoated wells (nonspecific background) from the average OD_450nm_ of duplicate hNephrin_G1059_ coated wells for each individual patient sample. A relative antibody titer was then determined from a standard curve that was generated using a single positive patient sample with a 1:100 dilution defined as containing 1000 Units/ml. The threshold for anti-nephrin antibody positivity (187 Units/ml) was defined as the maximum antibody titer, as the cohort was not normally distributed, in a healthy control population (n=30) with no known kidney disease (dotted line) to maximize specificity. The earliest serum sample available during active disease (urine protein creatinine ratio (UPCR) > 3 g/g on the day of sample collection) was positive for anti-nephrin antibodies in 18 (29%) of 62 patients with biopsy proven MCD from the NEPTUNE cohort. 53 (98%) of 54 nephrotic control patients with anti-human phospholipase A_2_ receptor (hPLA_2_R+) antibodies, as determined by clinical ELISA and IIFT assays (Euroimmun), were negative for anti-nephrin antibodies. The intra and inter-assay coefficient of variances for the anti-nephrin antibody ELISA were 5.56% and 14.36% respectively. The antibody titer for the NEPTUNE patients and controls are given in Supplemental Table S1. **(b)** 11 of the 18 NEPTUNE patients who were anti-nephrin antibody positive during active disease (blue bar) had a subsequent serum sample available during complete remission (UPCR < 0.3 g/g on the day of sample collection) which tested anti-nephrin antibody negative (red bar) in all cases. Dotted line indicates threshold for positive antibody titer (187 Units/ml). Student t-test was used to compare differences between the active and remission samples **p<0.01, ***p<0.001. **(c)** The same serum samples evaluated by ELISA from the NEPTUNE cohort (b) were evaluated for their ability to immunoprecipitate nephrin from HGE (derived from non-diseased human kidney). In keeping with the ELISA results, only serum obtained during active disease (indicated by an arrow with an A (blue colored) above it) immunoprecipitated nephrin, whereas serum obtained during remission (indicated by an arrow with a R (red colored) above it) did not. Total IgG was comparable between active and remission samples for each patient.

The patients’ clinical characteristics (Table S2) and the median time from enrollment to complete remission (CR) were similar between the anti-nephrin ab positive and negative groups (4.4 months vs 5.4 months respectively; p=0.7288) (Fig. S2). However, the relapse-free period was shorter for the anti-nephrin ab positive group compared with the ab negative group, although this finding did not reach conventional levels of statistical significance (median time to relapse 6.0 months vs 21.57 months respectively; p=0.0945) (Fig. S2).

A subsequent serum sample was available during either complete (UPCR < 0.3 g/g) or partial remission (>50% reduction in proteinuria) from 12 of the 18 anti-nephrin ab positive patients, in whom we observed a complete absence or a significant reduction of nephrin autoantibodies respectively (Fig. 1b, Figs. S3, S4). In keeping with the ELISA results, only serum obtained during active disease or partial remission immunoprecipitated nephrin from healthy donor kidney derived human glomerular extract (HGE), whereas serum obtained during complete remission did not (Fig. 1c).

To further investigate a potential pathogenic role of these nephrin autoantibodies, we next sought to establish whether they are present within kidneys of patients with MCD. One limitation of the NEPTUNE cohort is that biopsy material from these patients was not available for further evaluation and so we turned to our own institution and collaborators for biopsy and serum samples.

For many years, we have observed a delicate punctate staining for IgG in a subset of patients with MCD (MCD+) by routine immunofluorescence staining that is distinct from the background (Fig. S5a). It is much more subtle when compared to the prominent IgG staining observed in MN (Fig. S5a), and while this feature has been previously described^14^, its significance has not been fully established. We therefore hypothesized that this subtle IgG may represent autoantibodies targeting nephrin. To limit the possibility of staining artifacts, we routinely use a directly conjugated FITC anti-huma IgG F(ab)_2_ ab which we have independently validated with a distinct unconjugated anti-human IgG ab, an anti-light chain ab and isotype specific anti-IgG abs that all show an identical staining pattern (Fig. S6). Importantly, we observe a complete lack of concurrent glomerular albumin staining, indicating that this feature is IgG selective and does not reflect non-specific protein resorption (Fig. S6).

We utilized confocal microscopy to further evaluate this punctate IgG in renal biopsies that were received and processed by us over the last 3 years. We observed two predominant patterns of IgG distribution: GBM-associated fine punctate or curvilinear structures and more apically located punctate and vaguely vesicular clusters. These disparate staining patterns may reflect different stages of antibody binding and/or redistribution. In all the MCD+ biopsies evaluated, we observed specific co-localization of nephrin with the punctate IgG and not the background (Fig 2a, Fig. S5b, Fig. S7) which was further corroborated in the control biopsies lacking this punctate IgG (Fig. 2a, Fig. S5b). Antigen specificity was evidenced by a clear spatial association of the IgG with the SD-associated nephrin but not with the podocyte foot process associated synaptopodin by confocal microscopy (Fig. 2a,b, Fig. S5c) and by Super-Resolution Structured Illumination Microscopy (SR-SIM) which achieves an even higher spatial resolution^24^ (Fig. 2c,d, Fig. S8). Furthermore, in those biopsies exhibiting the granular redistribution of nephrin away from the SD, as previously described in MCD^20,21^, the IgG did not co-localize with the three intracellular podocyte specific proteins; synaptopodin (foot process associated), podocin (SD associated) and WT1 (nuclear) (Fig. S9).

**Figure 2:**
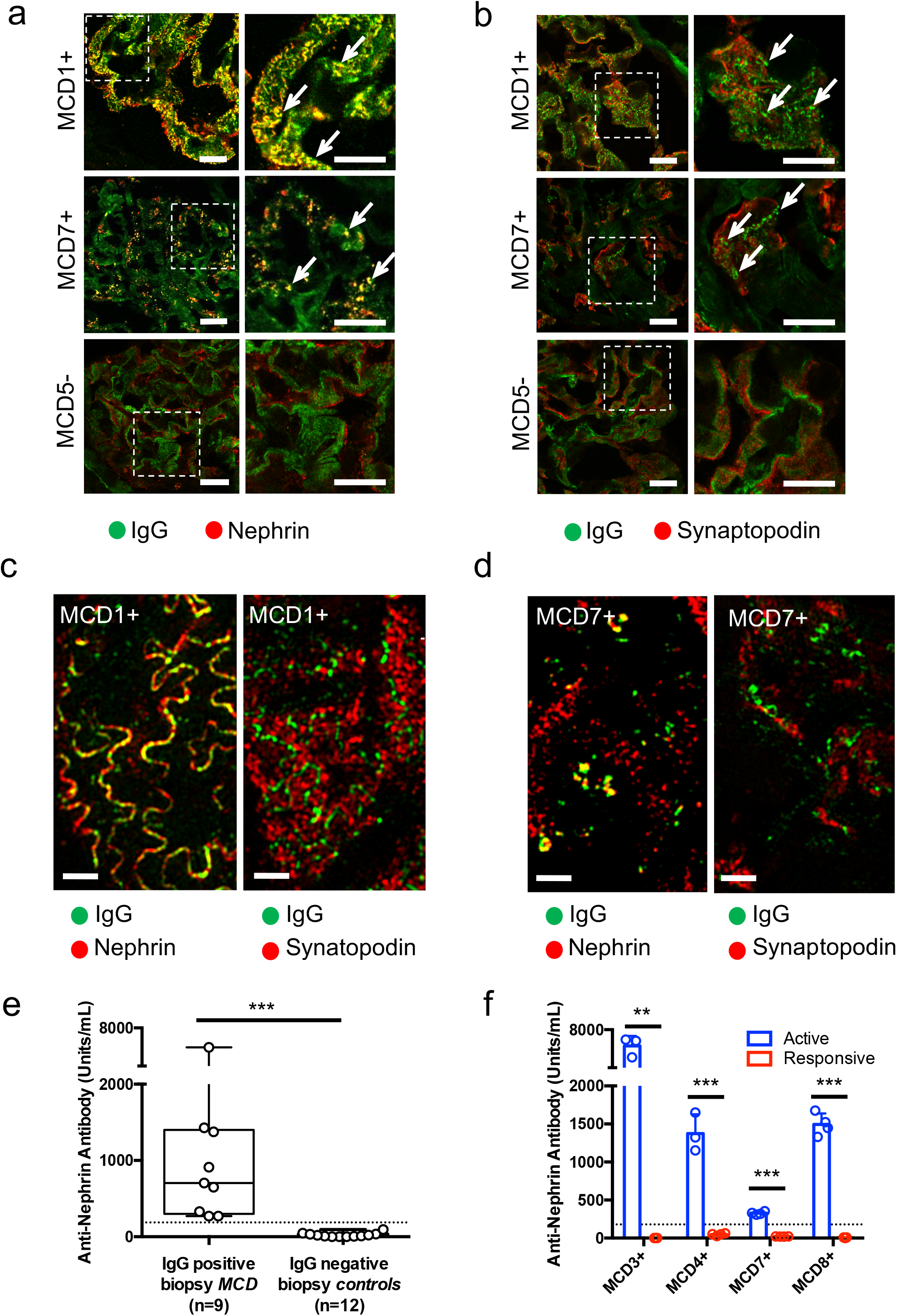
Renal biopsy imaging studies and serological testing for anti-nephrin antibodies in patients with biopsy proven MCD. **(a)** Representative confocal microscopy images of glomeruli in IgG-positive MCD (MCD1+/MCD7+) and IgG-negative MCD (MCD5-), stained for IgG (green) and the podocyte slit-diaphragm protein nephrin (red). There is a clear overlap (yellow) between IgG (green) and nephrin (red) specifically with the punctate IgG and not background as seen in the MCD+ biopsies (MCD1+/MCD7+) (white arrows) but not in MCD-biopsies. The right panels show magnified images of boxed areas of MCD1+ and MCD7+ biopsies. Scale bar: 10µm. **(b)** Representative confocal microscopy images of glomeruli in IgG-positive MCD (MCD1+/MCD7+) and IgG-negative MCD (MCD5-), stained for punctate IgG (green) (indicated with white arrow) and the cytoskeletal podocyte marker synaptopodin (red). There is no appreciable overlap between IgG and Synaptopodin in any of those cases. The right panels show magnified images of boxed areas of MCD1+ and MCD7+ biopsies. Scale bar: 10µm. **(c)** Super Resolution Structured Illumination Microscopy (SIM) images of 0.125 μm individual Z-slices showing *en face* views of the podocyte junction from a representative renal biopsy (MCD1+) in which the nephrin remains GBM-associated, forming a curvilinear pattern. The left image shows co-localization (yellow) of IgG (green) with the slit diaphragm protein nephrin (red), in contrast to mutual exclusivity with the foot process associated synaptopodin (red) shown in the right image, indicating intimate spatial association with nephrin along the podocyte slit diaphragm. (Full image stack shown in Figure S8). Scale bar: 1 µm. **(d)** SIM image of 0.125 µm individual Z-slices from a representative renal biopsy (patient MCD7+) in which nephrin is redistributed to a more granular pattern. The left image shows co-localization (yellow) of IgG (green) with the slit diaphragm protein nephrin (red), in contrast to mutual exclusivity with the foot process associated synaptopodin (red) shown in the right image, indicating a continued close spatial association of the IgG with the redistributed nephrin. (Full image stack shown in supplemental Figure S8). Scale bar: 1 µm. **(e)** All of the patients with MCD and IgG deposition on biopsy (n=9) were anti-nephrin antibody positive, whereas all of the control subjects lacking IgG deposition on biopsy, consisting of DN (n=2), Amyloidosis (n=1), IgG-negative FSGS (n=2), IgG-negative TL (n=1), normal (n=1), disease-free region of tumor nephrectomy (n=2) and IgG-negative MCD (n=3), were anti-nephrin antibody negative (n=12). The Mann-Whitney U test was used to compare differences between the groups ***p<0.001. **(f)** Serum/plasma samples were obtained from patients with biopsy proven IgG-positive MCD (MCD+) during active disease (within 7 days of presentation with NS) and follow-up samples were obtained during complete (MCD4+, MCD7+) or partial (MCD8+) remission on the day of sample collection. For MCD3+, the follow-up serum sample was obtained approximately 3 weeks after entering a period of sustained complete remission. The threshold for a positive anti-nephrin antibody titer of 187 U/ml (indicated by dotted line) was based on the upper limit of a healthy control population. Anti-nephrin antibodies in serum/plasma were undetectable or significantly reduced to below the threshold for positivity, (red bar) during clinical remission compared with those during active disease (blue bar). Complete remission was defined as urinary protein creatinine ratio (UPCR) < 0.3 g/g or urinary albumin creatinine ratio (UACR) < 0.2 g/g. Partial remission was defined as a > 50% reduction in proteinuria (UPCR) that did not fall below 0.3 g/g. Student t-test was used to compare differences between the active and remission samples **p<0.01, ***p<0.001.

To confirm that MCD+ patients with punctate IgG on renal biopsy do indeed have circulating autoantibodies against nephrin, we evaluated serum or plasma that was available specifically during active disease for 9 of them. As expected, all 9 patients were serologically positive for anti-nephrin antibodies by ELISA, in contrast to 12 control patients lacking punctate IgG on renal biopsy, who were all serologically negative (Fig. 2e, Table S3). A follow-up serum or plasma sample was available for 4 of the 9 MCD+ patients, which showed a significant reduction in antibody titer concordant with treatment response (Fig. 2f, Fig. S10). These findings were corroborated by IP (Fig. S11), and for all the patients in this study who were serologically positive for circulating nephrin autoantibodies they did not cross react with PLA_2_R (Fig. S12).

Finally, to highlight a potential role of pre-transplant nephrin autoantibodies in post-transplant disease recurrence, which generally shows morphologic features indistinguishable from MCNS, we identified a patient with childhood onset, steroid dependent MCNS and no underlying genetic basis (as determined by clinical whole exome sequencing) who progressed to ESKD requiring kidney transplantation. In keeping with a pathogenic role for anti-nephrin autoantibodies, the patient developed massive proteinuria early post-transplant, that in contrast to CNF^18,19^ was associated with high pre-transplant levels of nephrin autoantibodies (Fig. S13).

## Discussion

Herein, we describe the discovery of circulating autoantibodies against the extracellular domain of nephrin, the essential constituent of the podocyte SD, in a subset of patients with non-congenital acute nephrotic syndrome. These nephrin autoantibodies are specifically present in MCNS kidney biopsies, forming distinct clusters. Our observations share striking parallels with the autoimmune blistering skin condition pemphigus, in which circulating autoantibodies target the desmosomal cell adhesion molecules desmogleins (dsg)^22^. In pemphigus, these autoantibodies directly interfere with cell adhesion through redistribution, clustering and endocytosis of dsg that disrupts the integrity of the desmosome^22^. Previous reports of experimental anti-nephrin antibody mediated disruption of nephrin homophilic interactions further support this potential mechanism in MCNS^3^. Furthermore, pemphigus exhibits a rapid response to glucocorticoid treatment (within days to weeks) that cannot be explained by reduced IgG synthesis alone and may be due to compensatory desmoglein synthesis in keratinocytes^25^. Similarly, most cases of MCNS respond rapidly to glucocorticoids (within weeks) which have also been shown to upregulate nephrin cell surface expression in cultured human podocytes^26^. Based on these findings, we speculate that the IgG co-localizing with nephrin in MCNS kidney biopsies may represent *in situ* binding of nephrin autoantibodies. This binding may trigger the redistribution of nephrin and result in SD failure with acute onset, massive proteinuria.

Recurrent acute nephrotic syndrome in the allograft, referred to as “recurrent FSGS” (rFSGS) in patients with a history of acute nephrotic syndrome in the native kidney, is morphologically indistinguishable from MCNS. Therefore, we also included a study in a transplant patient with an initial diagnosis of MCD in the native kidney which then progressed to FSGS and eventually ESKD. This patient rapidly developed acute and high proteinuria post-transplant consistent with rFSGS, which was not confirmed by a biopsy; however, the pre-transplant serum showed high levels of anti-nephrin antibodies which decreased after aggressive and successful treatment with rituximab and plasmapheresis; this patient went into sustained remission and retained the transplant. We realize that without further corroboration in a larger rFSGS patient sample, no definitive conclusions can be drawn. However, this case illustrates the need to advocate for a comprehensive analysis of anti-nephrin antibodies also in this group of patients with a much more devastating prognosis.

In conclusion, our discovery adds new insights into the etiology of a disease that has been poorly understood for decades. Future studies will be needed to establish the prevalence of this new subset of anti-nephrin antibody positive NS and the prognostic benefit of anti-nephrin antibodies as a novel, minimally invasive biomarker. Nevertheless, our work provides a mechanistic rationale for consideration of B-cell targeted therapy in a subset of NS patients and allows us to molecularly define those patients who stand to benefit most from the development of new, targeted therapeutic strategies for anti-nephrin antibody positive NS.

## Methods

### Clinical samples (Kidney tissue and serum/plasma)

Renal biopsies were independently assessed by collaborating renal pathologists across four institutions: Brigham and Women’s Hospital (BWH), Massachusetts General Hospital (MGH), Boston Medical Center (BMC) and the Mayo Clinic. Serum/plasma was obtained from patients attending those institutions as either discarded samples originally collected for clinical analysis (BWH/BMC/Mayo clinic) or archival samples from the Kidney Disease Biobank (courtesy of Dr. Sushrut Waikar, Partners Healthcare, in accordance with Partners Healthcare IRB Approval for patients attending BWH or MGH and were consented for serum/plasma collection at the time of renal biopsy). Histological studies were performed on archival kidney tissue that was received for routine clinical evaluation and included diffuse podocytopathies, other nephrotic conditions, and non-neoplastic renal parenchyma from tumor nephrectomies. Medical record review, histological and serological studies were approved by the respective Institutional Review Boards (IRB) for those institutions. Similarly, sera were obtained from patients with biopsy proven minimal change disease (MCD) from the Nephrotic Syndrome Study Network (NEPTUNE) longitudinal study^23^ during active disease and where available, in remission. Healthy control sera were randomly selected from Partners Healthcare Biobank, specifically excluding those subjects with any renal or autoimmune disease. Sera from nephrotic patients were evaluated for anti-hPLA_2_R antibodies at the MGH Immunopathology laboratory using a commercial enzyme linked immunosorbent assay (ELISA) and indirect immunofluorescence test (IIFT) (Euroimmun). Samples were coded to preserve patient anonymity.

### Whole genome sequencing of the NEPTUNE cohort

Whole genome sequencing with a goal median depth of 30x was performed using Illumina Hi-seq. A standard pipeline, Gotcloud, was applied for sequence alignment and variant calling^27,28^. The variant analysis focused on approximately 70 genes implicated in Mendelian NS. To screen pathogenic variants in the 70 previously implicated Mendelian NS genes, we employed a pipeline similar to one that has been previously reported^29^. The pathogenicity variants were ultimately classified according to ACMG standards and guidelines^30^.

### Human glomerular extract

Human glomerular extract (HGE) was prepared as previously described by Beck *et al*^*31*^. Briefly, glomeruli were isolated from human kidneys deemed non-suitable for transplantation (that had been authorized for use in medical research) obtained from New England Donor Services, by graded sieving followed by isolation of glomerular proteins in RIPA buffer (Boston BioProducts). IgG was pre-cleared from tissue lysate by incubation with Protein G Plus agarose beads (Santa Cruz). Only kidneys with less than 20% global glomerulosclerosis, on routine wedge biopsy, were used for glomerular isolation.

### Routine renal biopsy processing

After biopsy acquisition, renal cortex was immediately allocated for light (10% neutral-buffered formalin), immunofluorescence (Zeus transport media) and electron microscopy (Karnovsky’s fixative) processing. For routine clinical immunofluorescence, 4 μm cryosections were fixed in 95% ethanol for 10 minutes and incubated with FITC-conjugated polyclonal rabbit F(ab)_2_ anti-human IgG antibody (Dako; F0315) diluted 1:20. FITC-conjugated sheep anti-human IgG1, IgG2, IgG3, IgG4 (Binding Site; AF006, AF007, AF008, AF009, respectively) diluted 1:20 were used for IgG subclass evaluation. Albumin was detected using FITC-conjugated polyclonal rabbit anti-human albumin (Dako; F0117) diluted 1:30. Sections were mounted using Dako fluorescence mounting medium (Dako; S3023) with a #1.5 coverslip. Immunofluorescence images were acquired on an Olympus BX53 microscope with an Olympus DP72 camera at 150 ms exposure.

### Confocal microscopy

For confocal microscopy, 4 μm cryosections of human kidney biopsies were fixed in 95% ethanol for 10 minutes and subsequently blocked for one hour at room temperature (RT) with phosphate buffer saline (PBS) supplemented with 0.2% fish gelatin, 2% bovine serum albumin (BSA) and 2% fetal bovine serum (FBS). All antibodies were diluted in this blocking solution and incubated for one hour at RT. Nephrin was detected using 1 μg/ml primary polyclonal sheep anti-human nephrin (R&D systems; AF4269) followed by a secondary AlexaFluor™ 568-conjugated donkey anti-sheep IgG (Invitrogen; A21099). Synaptopodin was detected using anti-synaptopodin (N-terminus) guinea pig polyclonal antiserum (Progen; GP94-N) diluted 1:1000 followed by a secondary AlexaFluor™ 568-conjugated goat anti-guinea pig IgG (Invitrogen; A11075) antibody. Podocin and Wilms Tumor 1 (WT1) were detected using a primary polyclonal rabbit anti-human podocin (Millipore Sigma; P0372) and a primary monoclonal rabbit anti-human WT1 clone SC06-41 (Invitrogen; MA5-32215) diluted 1:500 and 1:300 respectively, followed by a secondary AlexaFluor™ 568-conjugated donkey anti-rabbit IgG (Invitrogen; A10042). IgG immune deposits were detected using a primary monoclonal mouse anti-human IgG antibody (Abcam; ab200699) diluted 1:750 followed by a secondary AlexaFluor™ 488-conjugated donkey anti-mouse IgG (Invitrogen; A21202). All secondary AlexaFluor™-conjugated antibodies were diluted 1:500. Sections were mounted using Vectashield anti-fade mounting medium (Vectashield, H-1000) with a #1.5 coverslip and images were acquired on a Leica TCS SPE microscope.

### Structured Illumination Microscopy (SIM)

Structured Illumination Microscopy (SIM) imaging was performed on 4 μm fixed, frozen human kidney biopsy sections processed according to the aforementioned protocol for confocal microscopy. All images were collected using an OMX V4 Blaze (GE Healthcare) microscope equipped with three watercooled PCO.edge sCMOS cameras, 488 nm, 568 nm laser lines, and 528/48 nm, 609/37 nm emission filters (Omega Optical). Images were acquired with a 60X/1.42 Plan-Apochromat objective lens (Olympus) with a final pixel size of 80 nm. Z stacks of 4-8 μm, were acquired with a 0.125 μm z-spacing, and 15 raw images (three rotations with five phases each) were acquired per plane. Spherical aberration was minimized for each sample using immersion oil matching^24^. Super resolution images were computationally reconstructed from the raw data sets with a channel-specific, measured optical transfer function, and a Wiener filter constant of 0.001 using CUDA-accelerated 3D-SIM reconstruction code^32^. Axial and lateral chromatic misregistration was determined using a single biological calculation slide, prepared with human kidney tissue stained with a primary mouse anti-human IgG monoclonal antibody (Abcam; ab200699) followed by both secondary AlexaFluor™ 488-conjugated donkey anti-mouse IgG (Invitrogen; A21202) and AlexaFluor™ 568-conjugated goat anti-mouse IgG (Invitrogen; A11031) antibodies on the same tissue cryosection. Experimental data sets were then registered using the imwarp function in MATLAB (MathWorks) ^33^.

### Generation of recombinant human nephrin and phospholipase A_2_ receptor (PLA_2_R)

Separate plasmids encoding the extracellular subdomains of human nephrin (amino acids 1-1059), comprising the 8 Ig-like C2-type domains and a single fibronectin type III domain, and human phospholipase A2 receptor (hPLA_2_R), comprising the N-terminal ricin domain, fibronectin type II domain and 8 C-type lectin domains (CTLD), both with C-terminal polyhistidine (6XHIS) tags, were generated by standard cloning techniques. The correct sequences were confirmed by whole plasmid sequencing (MGH DNA core). HEK293-F cells (Thermo Fisher) were transfected with 0.5 μg plasmid per 10^6^ cells using 1.5 μg PEI (polyethylenimine). The plasmid and PEI were pre-incubated for 20 mins in Freestyle media (Thermo Fisher) at one tenth the final volume and then added dropwise to the cells. After 3-5 days, provided the cell viability was >95%, the cell culture media was harvested by centrifugation (300 xg for 10 mins). Imidazole was added to a final concentration of 10 mM and the media was filter sterilized (0.2 μm) on ice. Nickel NTA resin (Qiagen) was washed 3x with 10 mM Imidazole in PBS and then incubated with the filtered media overnight at 4°C on a roller mixer (Thermo Fisher). The Nickel NTA resin was then washed 3x with 10 mM Imidazole in PBS and the recombinant proteins were eluted with 300 mM Imidazole in PBS. The purity of the eluted fractions was confirmed by SDS-PAGE with a 4-12% Bis-Tris gel (Invitrogen), pooled together and concentrated to 1 ml using an Amnicon centrifugation filter with a 10K molecular weight cut off (Millipore). The resultant protein was run over a Sephadex^™^ 300 column and 0.5 ml fractions were collected. The purity of the eluted fractions was confirmed by SDS-PAGE on a 4-12% Bis-Tris gel (Invitrogen) and the concentration determined by measuring absorbance at 280 nm using a Nanodrop spectrophotometer (Thermo Fisher). Immunoreactivity of the purified nephrin was confirmed by Western blot analysis, under reducing conditions using a primary sheep anti-human nephrin antibody (R&D) followed by a secondary HRP-conjugated donkey anti-sheep IgG antibody (Jackson immunoresearch), and of the purified hPLA_2_R under non-reducing conditions using serum from a patient with known anti-PLA_2_R antibodies (determined by commercial ELISA and IIFT (Euroimmun)) diluted 1:1000 and a secondary HRP-conjugated donkey anti-human IgG antibody (Jackson Immunoresearch).

### Enzyme Linked Immunosorbent Assay (ELISA)

Please contact the corresponding author for information.

### Immunoprecipitation and Western Blot

Please contact the corresponding author for information.

## Data Availability

The data referred to in this manuscript are available upon request from the corresponding author.

## Acknowledgements

We acknowledge Lin Shao (Yale University) for CUDA-accelerated 3D-SIM reconstruction code, and Talley Lambert, Anna Payne-Jost and Jennifer Waters (Nikon Imaging Core, Harvard Medical School) for assistance with SIM acquisition. We are grateful to Dr. Opeyemi Olabisi for ApoL1 genotyping, and Drs. Emily Dulude, Durga Rao and Andrew Bentall for providing supporting clinical data. We are thankful to Dr. Colin Garvie and Dr. Kasia Handing for assistance with protein purification. We thank Terri Woo, Colleen Ford, Kristie Swett, Hui Chen and Brant Douglas for expert technical assistance, and Dr. Andrew Lichtman, Dr. David Salant, Dr. Moran Dvela, Dr. Juanchi Pablo, Dr. Silvana Bazua Valenti, Dr. Katherine Vernon, Dr. Valeria Padovano and Morgan Thompson for enlightening discussions. We thank Dr. Camden Bay from Harvard Catalyst for biostatistics consultation. A.J.B.W. was supported by T32 training fellowships T32HL007627 and T32DK007527. A.W. was supported by an Eleanor Miles Shore Fellowship by Harvard Medical School and is a current recipient of a NephCure/NEPTUNE ancillary study award. G.L. is supported by NIH grant DK007053-45S1. A.G. is supported by NIH grants DK099465 and DK095045. S.W. is supported by NIH grants U01DK085660, U01DK104308, and UG3DK114915. M.S. is supported by NIH grant R01DK119380 and R01DK1088085. A.G. has financial interest in Goldfinch Biopharma, which was reviewed and is managed by Brigham and Women’s Hospital, Mass General Brigham, the Broad Institute and Harvard University in accordance with their conflict of interest policies. A.J.B.W., K.H.K., A.W., J.M.H and H.G.R. have filed a provisional patent “Methods for identifying and treating patients with antibody-mediated acquired primary or recurrent idiopathic nephrotic syndrome”. The Nephrotic Syndrome Study Network Consortium (NEPTUNE), U54-DK-083912, is a part of the National Institutes of Health (NIH) Rare Disease Clinical Research Network (RDCRN), supported through a collaboration between the Office of Rare Diseases Research (ORDR), NCATS, and the National Institute of Diabetes, Digestive, and Kidney Diseases. Additional funding and/or programmatic support for this project has also been provided by the University of Michigan, the NephCure Kidney International and the Halpin Foundation.

