## Supplemental figures and tables for "Autoantibodies against nephrin elucidate a novel autoimmune phenomenon in proteinuric kidney disease"

### SUPPLEMENTARY APPENDIX

#### Table of Contents


| | |
|---|---|
| Table S1 | 2 |
| Figure S1 | 6 |
| Table S2 | 7 |
| Figure S2 | 8 |
| Figure S3 | 9 |
| Figure S4 | 11 |
| Figure S5 | 12 |
| Figure S6 | 14 |
| Figure S7 | 15 |
| Figure S8 | 16 |
| Figure S9 | 17 |
| Table S3 | 18 |
| Figure S10 | 19 |
| Figure S11 | 20 |
| Figure S12 | 21 |
| Clinical case details | 22 |
| Figure S13 | 23 |

*NEPTUNE cohort*

| Patient | Age (5yr range) | Sex | Race/ Ethnicity | UPCR (g/g) | Peak sCr (mg/dL) | α–Nephrin Ab (U/ml) | Follow up (months) | Remission | Relapse | Treatment |
|---|---|---|---|---|---|---|---|---|---|---|
| N1 | 5 | Male | White* | 25.70 | 0.5 | 2864 ± 832 | 53 | Partial | N/A | P/CNI/RIT/MMF |
| N2 | 55 | Male | Black | 7.04 | 4.53 | 1271 ± 101 | 23 | Complete | Yes | P/CNI/MMF |
| N3 | 20 | Male | White | 8.68 | 0.9 | 963 ± 72 | 10 | Complete | No | P/CNI/RIT |
| N4 | 15 | Female | White | 12.06 | 5.45 | 745 ± 99 | 43 | Complete | No | P/MMF |
| N5 | 40 | Male | Asian | 19.00 | 1.29 | 736 ± 26 | 29 | Complete | Yes | P |
| N6 | 35 | Male | Asian | 27.49 | 1.24 | 512 ± 48 | 38 | Complete | Yes | P |
| N7 | 25 | Female | Black | 5.78 | 1.25 | 442 ± 62 | 31 | Complete | No | P/MMF |
| N8 | 50 | Female | White* | 19.93 | 2.52 | 423 ± 53 | 1 | Complete | No | P |
| N9 | 5 | Male | Black | 10.58 | 0.41 | 392 ± 62 | 28 | Complete | Yes | P/CNI |
| N10 | 20 | Male | White* | 3.43 | 0.88 | 381 ± 38 | 18 | Complete | No | P |
| N11 | 60 | Female | White* | 11.22 | 2.6 | 320 ± 67 | 31 | Complete | Yes | P/RIT |
| N12 | 5 | Male | Asian | 4.28 | 0.83 | 305 ± 115 | 56 | Partial | N/A | P/CNI/MMF |
| N13 | 5 | Male | Asian | 9.60[#] | 0.34 | 293 ± 57 | 14 | Complete | Yes | P/CNI/RIT |
| N14 | 40 | Female | White | 7.77 | 0.8 | 273 ± 135 | 6 | Complete | Yes | P/MMF |
| N15 | 20 | Male | Asian | 9.57 | 0.84 | 253 ± 62 | 60 | Complete | Yes | P/CNI |
| N16 | 5 | Male | Multi | 3.49 | 0.3 | 235 ± 19 | 12 | Partial | N/A | P/CNI/MMF |
| N17 | 15 | Male | Black | 7.85 | 0.93 | 193 ± 22 | 38 | Complete | Yes | P/MMF |
| N18 | 30 | Female | White* | 3.54 | 0.6 | 191 ± 71 | 19 | Complete | No | P/MMF |
| N19 | 10 | Male | White | 4.80 | 0.64 | 172 ± 18 | 33 | Complete | No | P/CNI/MMF |
| N20 | 50 | Female | White | 8.42 | 0.91 | 172 ± 32 | 3 | Complete | Yes | P/MMF |
| N21 | 20 | Male | Black | 8.41 | 0.7 | 165 ± 35 | 80 | Complete | Yes | P/CNI |
| N22 | 25 | Male | Black | 6.17 | 3.16 | 156 ± 30 | 21 | Complete | No | P/CNI |
| N23 | 10 | Female | Black | 10.38 | 10 | 137 ± 15 | 57 | Complete | Yes | P/CNI/MMF |
| N24 | 15 | Female | Black | 6.66 | 1.02 | 134 ± 69 | 18 | Complete | Yes | P |
| N25 | 55 | Male | White | 3.28 | 1.10 | 129 ± 76 | 55 | Complete | No | CTX/RIT/FLU |
| N26 | 5 | Female | Multi | 4.71 | 0.52 | 129 ± 48 | 53 | Complete | Yes | P/CNI |
| N27 | 15 | Male | White | 7.34 | 0.86 | 117 ± 18 | 30 | Complete | Yes | P/CNI/RIT |
| N28 | 5 | Female | White | 7.58 | 0.51 | 116 ± 36 | 60 | Complete | No | P/CTX/CNI |
| N29 | 30 | Male | White | 13.84 | 0.7 | 98 ± 80 | 47 | Complete | No | P |
| N30 | 5 | Male | Black | 16.24 | 0.40 | 86 ± 40 | 48 | Complete | Yes | P/CNI |
| N31 | 15 | Male | Asian | 10.03 | 0.88 | 85 ± 45 | 25 | Complete | Yes | P/CNI |
| N32 | 35 | Female | White* | 3.00 | 0.71 | 82 ± 18 | 20 | Complete | No | P |
| N33 | 5 | Male | White* | 20.93 | 0.32 | 80 ± 11 | 20 | Complete | Yes | P/CNI/RIT |
| N34 | 20 | Male | White | 5.25 | 0.8 | 78 ± 12 | 30 | Complete | Yes | P/MMF/RIT |
| N35 | 20 | Female | Black | 5.18 | 0.7 | 74 ± 54 | 13 | Complete | No | P/CNI |
| N36 | 5 | Female | Unknown * | 10.51 | 0.5 | 72 ± 85 | 54 | Complete | Yes | P/CTX |
| N37 | 5 | Female | Black | 40.38 | 0.33 | 71 ± 110 | 28 | Complete | No | P/CNI |
| N38 | 15 | Female | Black | 4.71 | 1.1 | 70 ± 12 | 57 | Complete | No | P/MMF |
| N39 | 10 | Male | Multi | 5.54 | 1.82 | 59 ± 40 | 53 | Complete | Yes | P/CNI |
| N40 | 20 | Male | White | 7.66 | 0.91 | 55 ± 5 | 25 | Complete | No | P |
| N41 | 15 | Female | White | 12.53 | 0.57 | 52 ± 51 | 23 | Complete | No | P |
| N42 | 60 | Female | Black | 3.46 | 1.77 | 38 ± 30 | 13 | Complete | Yes | P/CNI |

| | | | | | | | | | | |
|---|---|---|---|---|---|---|---|---|---|---|
| N43 | 15 | Male | White* | 3.52 | 0.59 | 35 ± 12 | 46 | Complete | No | P/CNI/MMF |
| N44 | 10 | Female | Asian | 5.18 | 0.47 | 33 ± 13 | 37 | Partial | N/A | P/CNI/MMF |
| N45 | 15 | Female | Unknown * | 3.07 | 1.2 | 31 ± 31 | 47 | Complete | No | P |
| N46 | 15 | Male | White | 3.01 | 0.9 | 29 ± 58 | 60 | Complete | Yes | P |
| N47 | 40 | Male | White* | 6.94 | 3.62 | 25 ± 30 | 55 | Complete | No | P |
| N48 | 5 | Female | White* | 12.70 | 0.5 | 21 ± 18 | 54 | Complete | Yes | P/CNI/MMF |
| N49 | 15 | Female | White | 8.34 | 0.61 | 19 ± 14 | 3 | None | N/A | P/CNI/MMF |
| N50 | 10 | Female | White | 17.92 | 0.42 | 18 ± 19 | 54 | None | N/A | P/CNI |
| N51 | 10 | Male | Black | 12.14 | 0.6 | 15 ± 13 | 51 | Complete | Yes | P/MMF/CNI |
| N52 | 20 | Female | White* | 11.31 | 0.83 | 12 ± 21 | 36 | Complete | No | P |
| N53 | 5 | Male | Black | 10.32 | 0.7 | 10 ± 14 | 57 | Complete | Yes | P/CNI |
| N54 | 10 | Male | White | 9.70 | 0.52 | 9 ± 8 | 26 | Complete | Yes | P/CNI |
| N55 | 5 | Female | Black | 8.89 | 0.58 | 7 ± 12 | 51 | Complete | Yes | P/CNI |
| N56 | 30 | Female | White* | 7.12 | 1.06 | 6 ± 8 | 55 | Partial | N/A | P/CNI/MMF |
| N57 | 5 | Male | Multi | 8.94 | 0.4 | 6 ± 7 | 52 | Complete | Yes | P/CNI/MMF/CTX |
| N58 | 10 | Male | White | 6.48 | 0.6 | 3 ± 8 | 32 | Complete | Yes | P/MMF |
| N59 | 5 | Female | White | 9.69 | 0.46 | 0 ± 0 | 52 | Complete | Yes | P/CNI |
| N60 | 10 | Male | White | 5.64 | 0.6 | 0 ± 0 | 53 | Complete | Yes | P/MMF |
| N61 | 50 | Male | White | 5.63 | 1.11 | 0 ± 0 | 43 | Complete | Yes | P/RIT |
| N62 | 15 | Male | Multi | 10.76 | 1.01 | 0 ± 0 | 27 | Complete | Yes | P/CNI |

***Healthy Controls***

| **Patient** | **Age (5yr range)** | **Sex** | **Race** | **α–Nephrin Ab (U/ml)** |
|---|---|---|---|---|
| CNT1 | 70 | Male | White | 187 ± 45 |
| CNT2 | 30 | Female | White | 174 ± 84 |
| CNT3 | 25 | Male | White | 100 ± 60 |
| CNT4 | 65 | Female | White | 57 ± 35 |
| CNT5 | 55 | Female | White | 49 ± 11 |
| CNT6 | 20 | Male | White | 43 ± 21 |
| CNT7 | 65 | Male | White | 41 ± 28 |
| CNT8 | 50 | Male | White | 41 ± 34 |
| CNT9 | 35 | Male | White | 39 ± 28 |
| CNT10 | 60 | Female | White | 36 ± 52 |
| CNT11 | 45 | Female | White | 33 ± 35 |
| CNT12 | 80 | Female | White | 27 ± 26 |
| CNT13 | 80 | Male | White | 23 ± 34 |
| CNT14 | 60 | Male | White | 19 ± 14 |
| CNT15 | 40 | Male | White | 18 ± 19 |
| CNT16 | 60 | Male | White | 16 ± 13 |
| CNT17 | 20 | Male | White | 12 ± 15 |
| CNT18 | 20 | Male | White | 12 ± 11 |
| CNT19 | 20 | Male | White | 8 ± 8 |
| CNT20 | 60 | Female | White | 6 ± 8 |
| CNT21 | 50 | Male | White | 6 ± 8 |
| CNT22 | 10 | Female | White | 6 ± 11 |
| CNT23 | 45 | Male | White | 4 ± 9 |

| | | | | |
|---|---|---|---|---|
| CNT24 | 55 | Female | White | 3 ± 7 |
| CNT25 | 40 | Male | White | 0 ± 0 |
| CNT26 | 65 | Male | White | 0 ± 0 |
| CNT27 | 50 | Male | White | 0 ± 0 |
| CNT28 | 10 | Male | White | 0 ± 0 |
| CNT29 | 30 | Male | White | 0 ± 0 |
| CNT30 | 20 | Male | White | 0 ± 0 |

***h$PLA_2R$+ cohort***

| **Patient** | **Age (5yr range)** | **Gender** | **$\alpha$–Nephrin Ab (U/ml)** |
|---|---|---|---|
| $PLA_2R1$ | 75 | Female | 196 ± 89 |
| $PLA_2R2$ | 70 | Female | 184 ± 119 |
| $PLA_2R3$ | 70 | Male | 162 ± 141 |
| $PLA_2R4$ | 65 | Female | 160 ± 71 |
| $PLA_2R5$ | 75 | Female | 132 ± 101 |
| $PLA_2R6$ | 70 | Female | 128 ± 83 |
| $PLA_2R7$ | 70 | Male | 101 ± 80 |
| $PLA_2R8$ | 75 | Male | 80 ± 22 |
| $PLA_2R9$ | 40 | Male | 61 ± 13 |
| $PLA_2R10$ | 70 | Male | 53 ± 61 |
| $PLA_2R11$ | 85 | Male | 52 ± 28 |
| $PLA_2R12$ | 40 | Male | 45 ± 16 |
| $PLA_2R13$ | 70 | Male | 42 ± 14 |
| $PLA_2R14$ | 80 | Female | 37 ± 57 |
| $PLA_2R15$ | 60 | Male | 36 ± 30 |
| $PLA_2R16$ | 55 | Female | 30 ± 45 |
| $PLA_2R17$ | 30 | Male | 27 ± 13 |
| $PLA_2R18$ | 55 | Male | 25 ± 44 |
| $PLA_2R19$ | 50 | Male | 21 ± 8 |
| $PLA_2R20$ | 60 | Male | 20 ± 17 |
| $PLA_2R21$ | 60 | Male | 20 ± 30 |
| $PLA_2R22$ | 50 | Male | 17 ± 14 |
| $PLA_2R23$ | 40 | Female | 10 ± 7 |
| $PLA_2R24$ | 40 | Female | 10 ± 17 |
| $PLA_2R25$ | 40 | Male | 10 ± 9 |
| $PLA_2R26$ | 60 | Male | 10 ± 17 |
| $PLA_2R27$ | 70 | Male | 9 ± 16 |
| $PLA_2R28$ | 45 | Female | 6 ± 8 |
| $PLA_2R29$ | 70 | Male | 5 ± 9 |
| $PLA_2R30$ | 60 | Female | 5 ± 8 |
| $PLA_2R31$ | 45 | Male | 4 ± 7 |
| $PLA_2R32$ | 60 | Male | 3 ± 6 |
| $PLA_2R33$ | 60 | Male | 2 ± 4 |
| $PLA_2R34$ | 85 | Female | 2 ± 4 |
| $PLA_2R35$ | 80 | Male | 2 ± 3 |
| $PLA_2R36$ | 70 | Male | 1 ± 2 |
| $PLA_2R37$ | 65 | Male | 0 ± 0 |

| | | | |
|---|---|---|---|
| $PLA_2R38$ | 65 | Male | 0 ± 0 |
| $PLA_2R39$ | 55 | Female | 0 ± 0 |
| $PLA_2R40$ | 40 | Female | 0 ± 0 |
| $PLA_2R41$ | 60 | Female | 0 ± 0 |
| $PLA_2R42$ | 60 | Female | 0 ± 0 |
| $PLA_2R43$ | 60 | Male | 0 ± 0 |
| $PLA_2R44$ | 55 | Female | 0 ±0 |
| $PLA_2R45$ | 45 | Male | 0 ± 0 |
| $PLA_2R46$ | 75 | Male | 0 ± 0 |
| $PLA_2R47$ | 85 | Female | 0 ± 0 |
| $PLA_2R48$ | 65 | Male | 0 ± 0 |
| $PLA_2R49$ | 85 | Female | 0 ± 0 |
| $PLA_2R50$ | 55 | Female | 0 ± 0 |
| $PLA_2R51$ | 55 | Female | 0 ± 0 |
| $PLA_2R52$ | 75 | Male | 0 ± 0 |
| $PLA_2R53$ | 90 | Male | 0 ± 0 |
| $PLA_2R54$ | 80 | Male | 0 ± 0 |

**Table S1. Clinical information for the NEPTUNE patients and controls evaluated for nephrin autoantibodies.** The table provides relevant clinical information for the patients or controls. All patients in the NEPTUNE cohort had biopsy proven minimal change disease (MCD); however, the renal biopsy IgG deposition status was not reported and neither immunofluorescence images nor biopsy material were available for further assessment. Proteinuria values (Urine Protein Creatinine ratio (UPCR)) are from the same day (or within one day) that the serum sample was collected for anti-nephrin antibody ($\alpha$-Nephrin Ab) testing during active disease. [#]For patient N13, the UPCR was calculated to be 323 g/g on the day of serum collection and so the value for the next available UPCR (assessed 20 days later) is given. Peak sCr (serum creatinine) was the highest serum creatinine reached during the follow-up period. Partial remission was defined as > 50% reduction in the UPCR and complete remission (CR) as UPCR < 0.3 g/g. A patient was deemed to have relapsed with a UPCR > 3 g/g after first reaching CR. In those patients not reaching CR, the relapse status is not applicable (N/A). Serum was obtained from a randomly selected healthy control cohort from Partners Healthcare Biobank. The threshold for a positive anti-nephrin antibody titer was based on the maximum value for the healthy cohort of 187 U/ml. Antibody titer is given as the mean ± S.D. of replicate samples (n≥3) for each patient. Serum from patients who tested positive for anti-human $PLA_2R$ antibodies (h$PLA_2R$+) by two clinically validated assays, ELISA and IIFT (Euroimmun), were obtained from MGH Immunopathology Laboratory. Designations: P, prednisolone; CNI, calcineurin inhibitor; MMF, mycophenolate mofetil; CTX, cyclophosphamide; RIT, rituximab; FLU, flucytosine; * indicates Hispanic or Latino ethnicity.

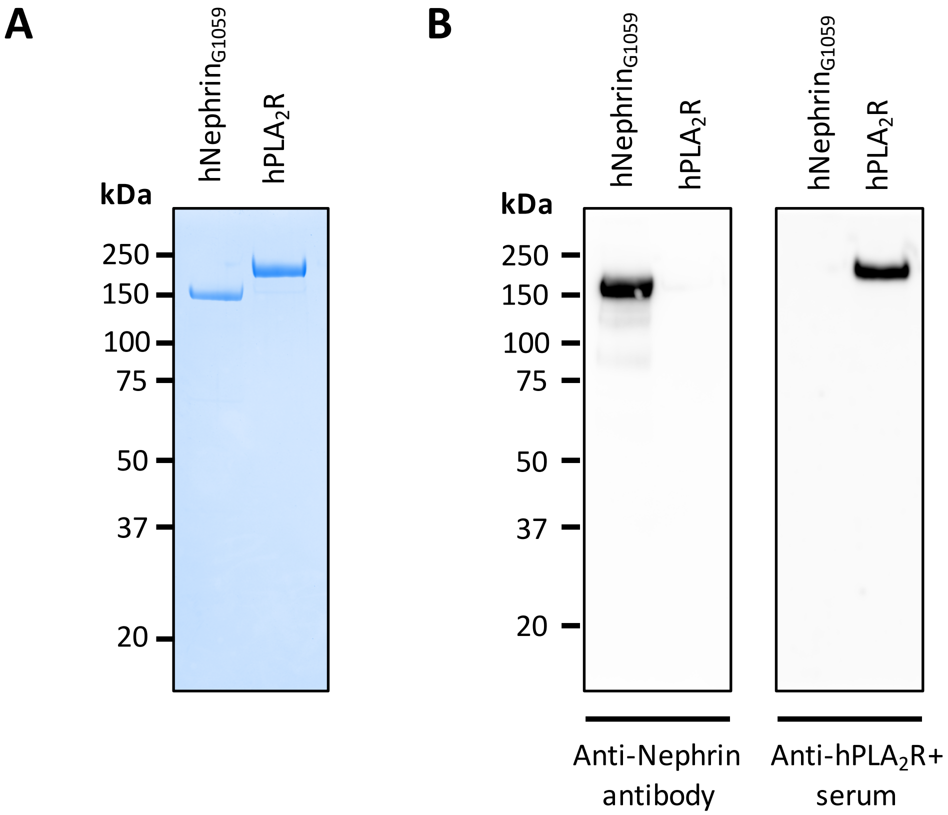


**Figure S1. Generation and validation of purified recombinant extracellular domains of human nephrin and human phospholipase $A_2$ receptor (h$PLA_2$R)**. Recombinant human extracellular domain of nephrin ($hNephrin_{G1059}$) (~150 kDa) and h$PLA_2$R (~180kDa) both with C-terminal polyhistidine (6XHIS) tags were affinity purified with Nickel-NTA resin followed by size exclusion chromatography. (A) Purity was confirmed by Coomassie staining of 1 µg recombinant protein resolved by SDS-PAGE under reducing conditions. (B) Immunoreactivity of (Left) 0.5 µg nephrin, under reducing conditions, was confirmed using a primary sheep anti-human nephrin antibody and (Right) 0.5 µg h$PLA_2$R, under non-reducing conditions, using serum from a patient with anti-h$PLA_2$R antibodies diluted 1:1000. The appropriate HRP-conjugated donkey anti-sheep or anti-human IgG secondary antibody was used for detection.

| | Anti-nephrin Ab Positive | Anti-nephrin Ab Negative | P value |
|---|---|---|---|
| Cases – no. (% of total cases) | 18 (29) | 44 (71) | |
| Age – yr | 20 (5.5-37.5) | 14 (6.25-17.75) | 0.11 |
| Male – no. (%) | 12 (67) | 23 (52) | 0.40 |
| Female – no. (%) | 6 (33) | 21 (48) | 0.40 |
| Disease onset | | | |
| Adulthood – no. (%) | 9 (50) | 12 (27) | 0.14 |
| Childhood (< 18 years) – no. (%) | 9 (50) | 32 (73) | 0.14 |
| Race | | | |
| White – no. (%) | 8 (44) | 24 (55) | 0.58 |
| Black – no. (%) | 4 (22) | 12 (27) | 0.76 |
| Asian – no. (%) | 5 (28) | 2 (4.5) | 0.018* |
| Multi – no. (%) | 1 (6) | 4 (9) | 1.00 |
| Unknown – no. (%) | 0 (0) | 2 (4.5) | 1.00 |
| Hispanic/Latino – no. (%) | 5 (28) | 9 (20) | 0.51 |
| No remission – no. (%) | 0 (0) | 2 (4.5) | 1.00 |
| Partial remission – no. (%) | 3 (17) | 2 (4.5) | 0.14 |
| Complete remission (CR) – no. (%) | 15 (83) | 40 (91) | 0.40 |
| Relapse after CR – no. (% of CR) | 9 (60) | 25 (63) | 1.00 |
| UPCR (g/g) at $\alpha$-nephrin Ab assay | 9.1 (5.4-13.79) | 7.6 (5.20-10.48) | 0.34 |
| Peak Serum creatinine (mg/dL) | 0.89 (0.58-1.6) | 0.7 (0.52-1.02) | 0.17 |

**Table S2. Comparison of the clinical and demographic data of patients from the NEPTUNE cohort grouped based on anti-nephrin antibody status.** The threshold for a positive anti-nephrin antibody ($\alpha$-nephrin Ab) level was based on a randomly selected, healthy control population with no renal disease. Complete remission was defined as a UPCR < 0.3 g/g. Partial remission was defined as a > 50% reduction in proteinuria that did not fall below 0.3 g/g. The continuous variables are presented as median (interquartile range). Statistical analysis was performed using the Mann-Whitney test for continuous variables and Fisher's exact test for categorical variables (*p<0.05).

**A**

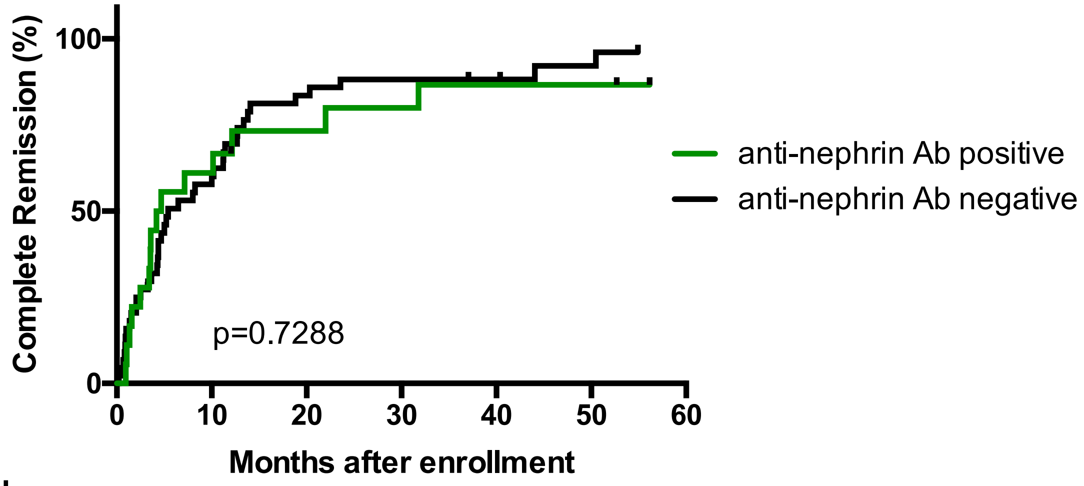


**No. at risk**

| | | | | | | | |
|---|---|---|---|---|---|---|---|
| Ab Positive | 18 | 7 | 4 | 3 | 2 | 2 | 0 |
| Ab Negative | 44 | 18 | 7 | 5 | 4 | 2 | 0 |

**B**

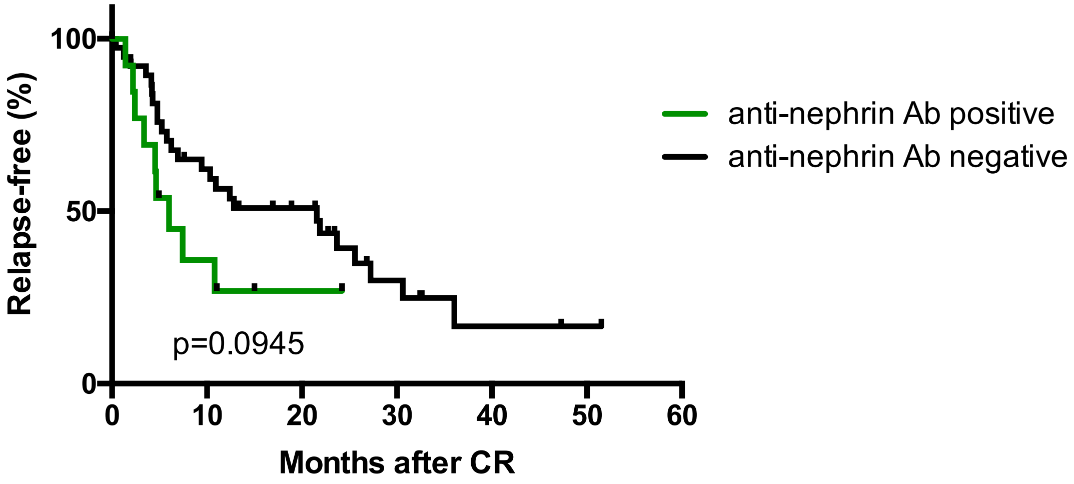


**No. at risk**

| | | | | | | | |
|---|---|---|---|---|---|---|---|
| Ab Positive | 15 | 4 | 1 | 0 | 0 | 0 | 0 |
| Ab Negative | 39 | 21 | 15 | 5 | 2 | 1 | 0 |

**Figure S2. Kaplan-Meier estimates for complete remission (CR) and relapse-free period in the NEPTUNE cohort.** (A) Time to complete remission from enrollment was similar between the anti-nephrin antibody (Ab) positive (median time 4.4 months) and negative groups (median time 5.4 months) by a log rank test (p=0.7288). (B) The relapse-free period following CR was shorter in the anti-nephrin antibody positive group (median time 6 months) compared to the antibody negative group (median time 21.57 months), although this finding did not reach statistical significance by a log rank test (p=0.0945). CR was defined as UPCR < 0.3 g/g. Relapse when UPCR > 3 g/g after reaching CR.

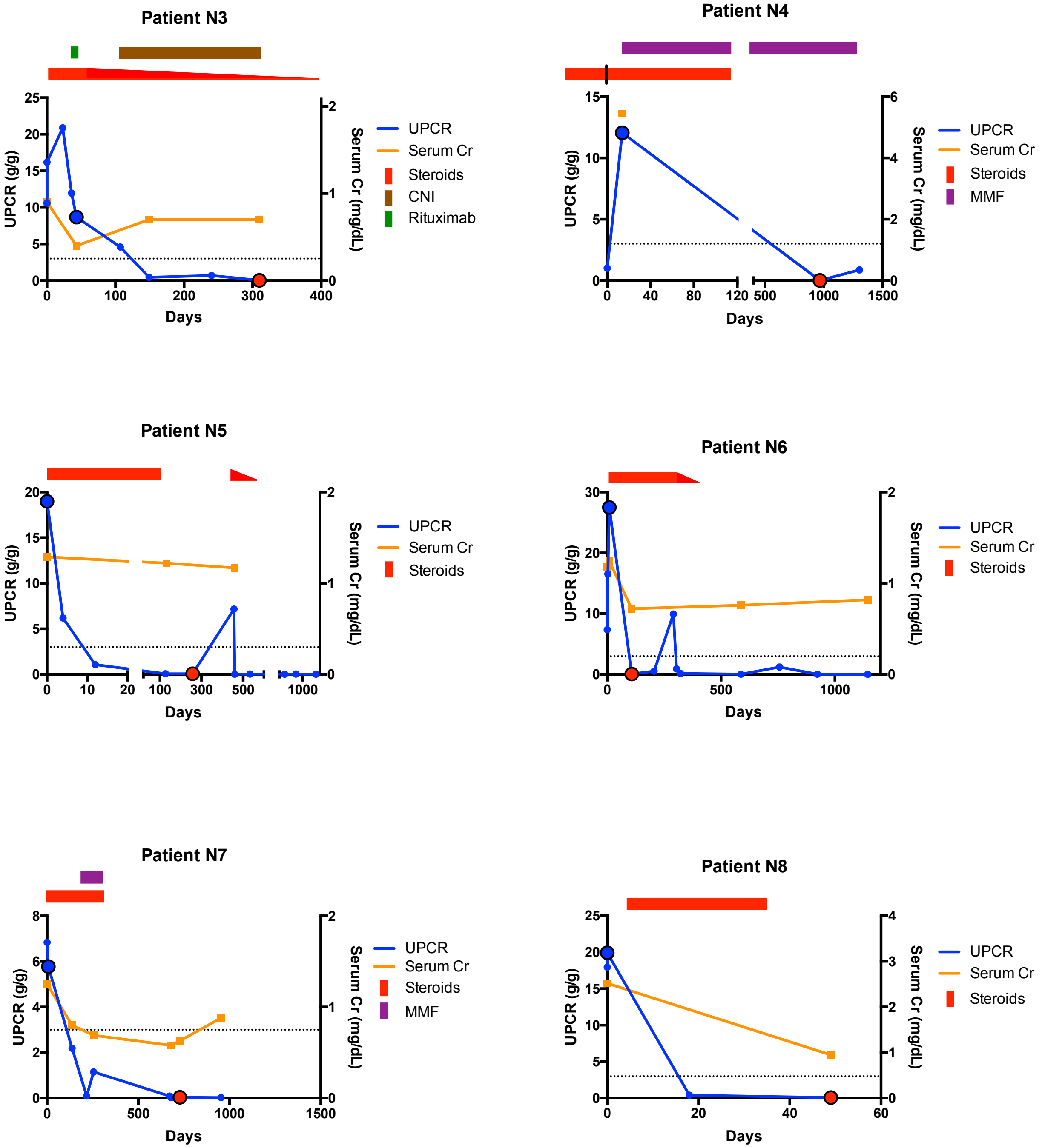

Patient N3
UPCR (g/g)
Serum Cr (mg/dL)
Days
UPCR
Serum Cr
Steroids
CNI
Rituximab
Patient N4
UPCR (g/g)
Serum Cr (mg/dL)
Days
UPCR
Serum Cr
Steroids
MMF
Patient N5
UPCR (g/g)
Serum Cr (mg/dL)
Days
UPCR
Serum Cr
Steroids
Patient N6
UPCR (g/g)
Serum Cr (mg/dL)
Days
UPCR
Serum Cr
Steroids
Patient N7
UPCR (g/g)
Serum Cr (mg/dL)
Days
UPCR
Serum Cr
Steroids
MMF
Patient N8
UPCR (g/g)
Serum Cr (mg/dL)
Days
UPCR
Serum Cr
Steroids

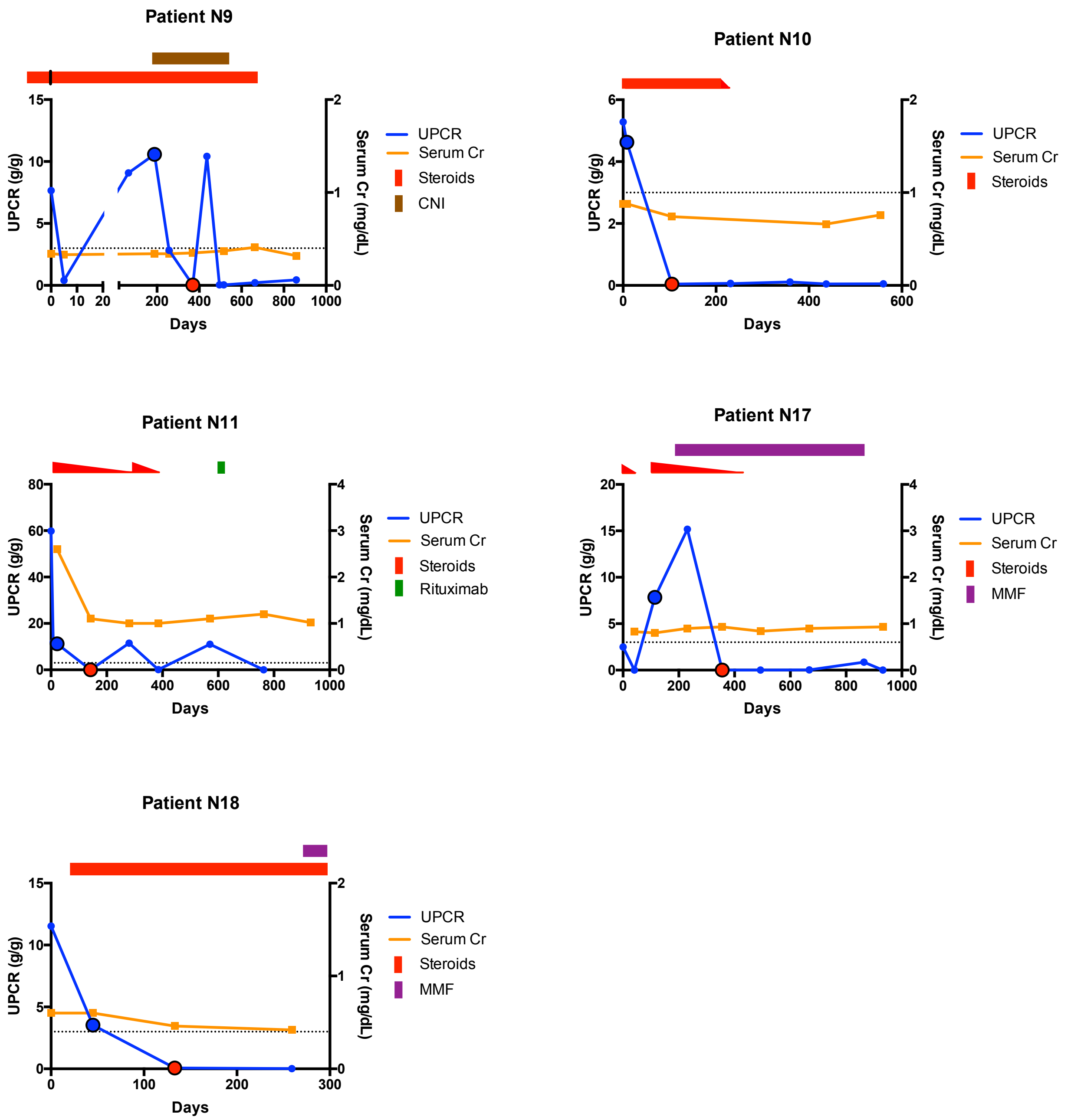


**Figure S3. The clinical course of patients from the NEPTUNE cohort with samples obtained during active disease and subsequent complete remission**. UPCR (g/g) and serum creatinine (Cr) are shown for each patient with serum samples available during active disease (large blue circle with black border) and during subsequent complete remission (large red circle with black border). The serum samples were evaluated for anti-nephrin antibodies and refer to the data shown in Figure 1B/C. Therapy is shown above each graph and for steroid treatment a dose reduction is indicated by a downward sloping wedge (red line). The dotted line indicates a UPCR of 3 g/g and complete remission was defined as UPCR < 0.3 g/g. Days, days following enrollment.

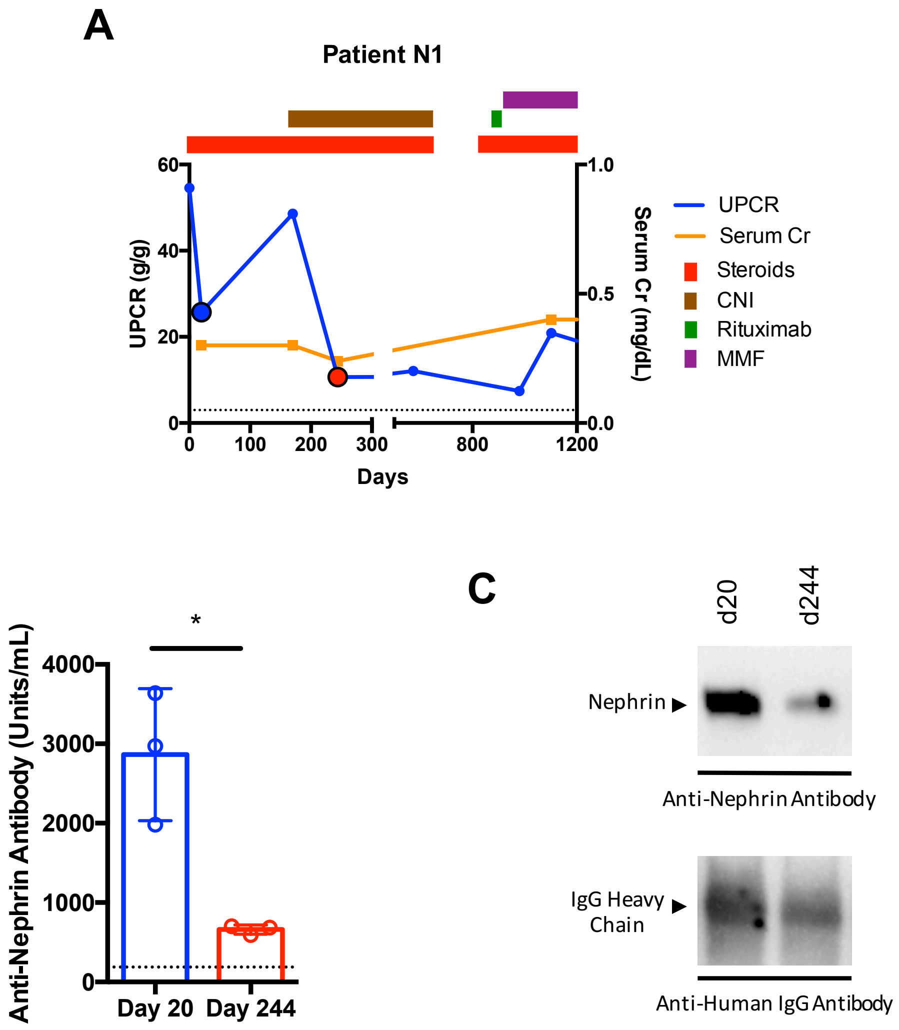


**Figure S4. Clinical and anti-nephrin antibody data for a single patient from the NEPTUNE cohort with active proteinuria and a partial response to therapy.** (A) Clinical course of patient with a serum sample available during active disease (large blue circle with black border) and partial remission (large red circle with black border) showing UPCR (g/g) (dotted line indicates UPCR of 3 g/g), serum creatinine (Cr) and treatment (shown above graph). (B) Significant reduction in anti-nephrin antibody titers associated with partial remission (*p=0.01) (dotted line indicates threshold for positive antibody titer of 187 U/ml based on a healthy control population). (C) The serum samples taken on day 20 (d20) and day 244 (d244) both immunoprecipitated nephrin from HGE derived from disease-free human kidneys, with the band intensity mirroring the antibody titers by ELISA. Days; days following enrollment. Partial remission was defined as a >50% reduction in UPCR between samples.

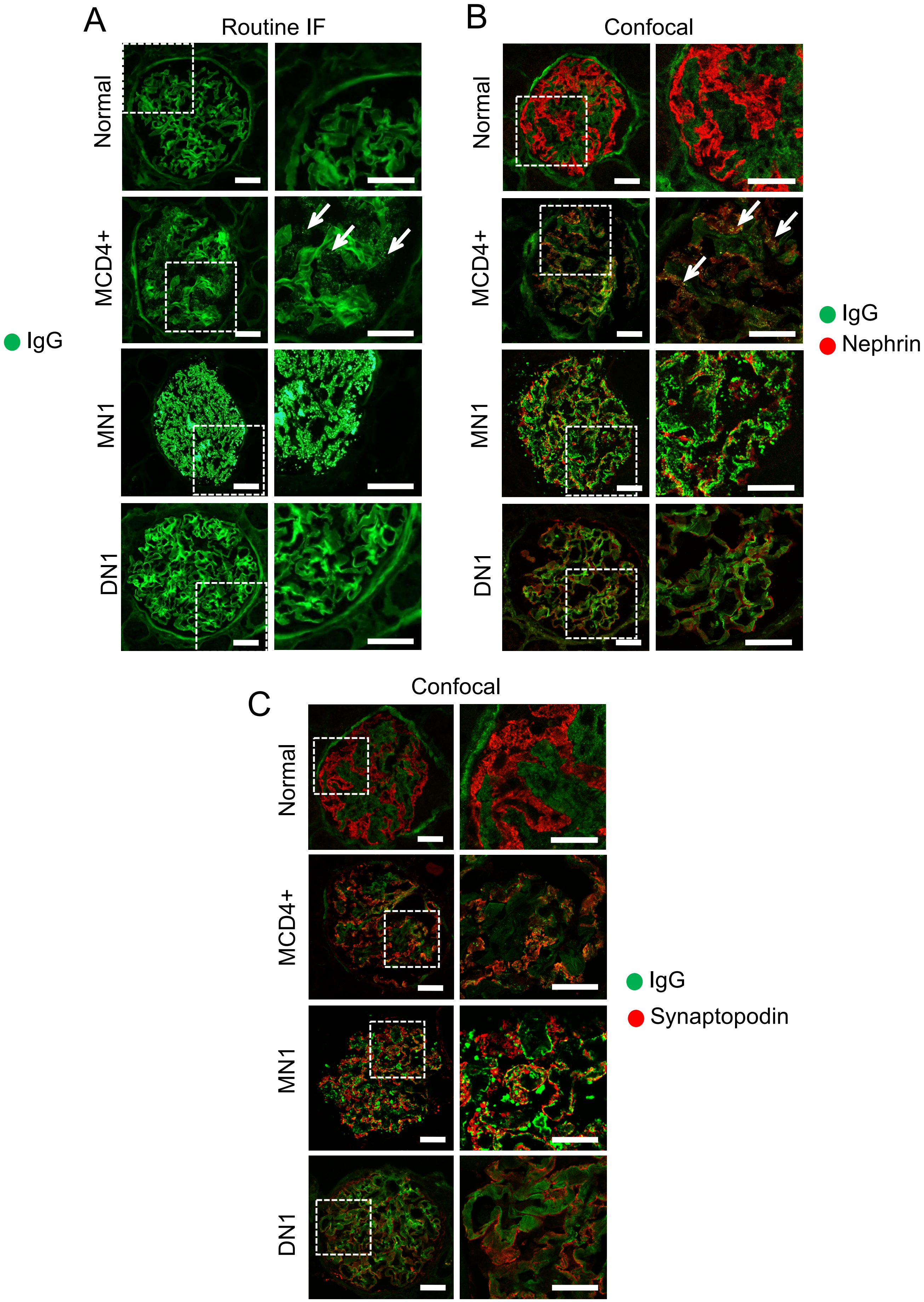
A
Routine IF
Normal
MCD4+
MN1
DN1
IgG
B
Confocal
Normal
MCD4+
MN1
DN1
IgG
Nephrin
C
Confocal
Normal
MCD4+
MN1
DN1
IgG
Synaptopodin

**Figure S5: Immunofluorescence microscopy images of IgG, IgG/Nephrin and IgG/Synaptopodin staining.** (a) Clinical epifluorescence images of glomeruli stained for IgG using FITC-conjugated anti-human IgG $(Fab)_2$ antibodies (ab). Right panel shows larger magnification of dotted square. A normal glomerulus shows low intensity linear staining along all basement membranes. In a subset of MCD (here shown MCD4+), a delicate punctate staining for IgG is observed in the extracapillary compartment, closely associated with GBMs (white arrows). In contrast, granular IgG staining in MN (here shown MN1) is of much stronger intensity. No granular or punctate staining is observed in other proteinuric conditions, including in patients with DN (here shown DN1). (b) Confocal microscopy images of the same cases stained for IgG and nephrin using primary unconjugated mouse anti-human IgG (green) and sheep anti-human nephrin (red) antibodies. Right panel shows larger magnification of dotted square. While reduction of nephrin staining intensity can be seen in all proteinuric conditions, co-localization of IgG with nephrin is only observed in MCD+ (white arrows). (c) Confocal microscopy images of same cases stained for IgG and synaptopodin using primary unconjugated mouse anti-human IgG (green) and guineapig anti-human synaptopodin (red) antibodies. Right panel shows larger magnification of dotted square. No appreciable co-localization of IgG staining with the intracellular actin-associated synaptopodin is seen in the normal kidney, MCD+, MN or DN. Scale bars: 20$\mu$m.

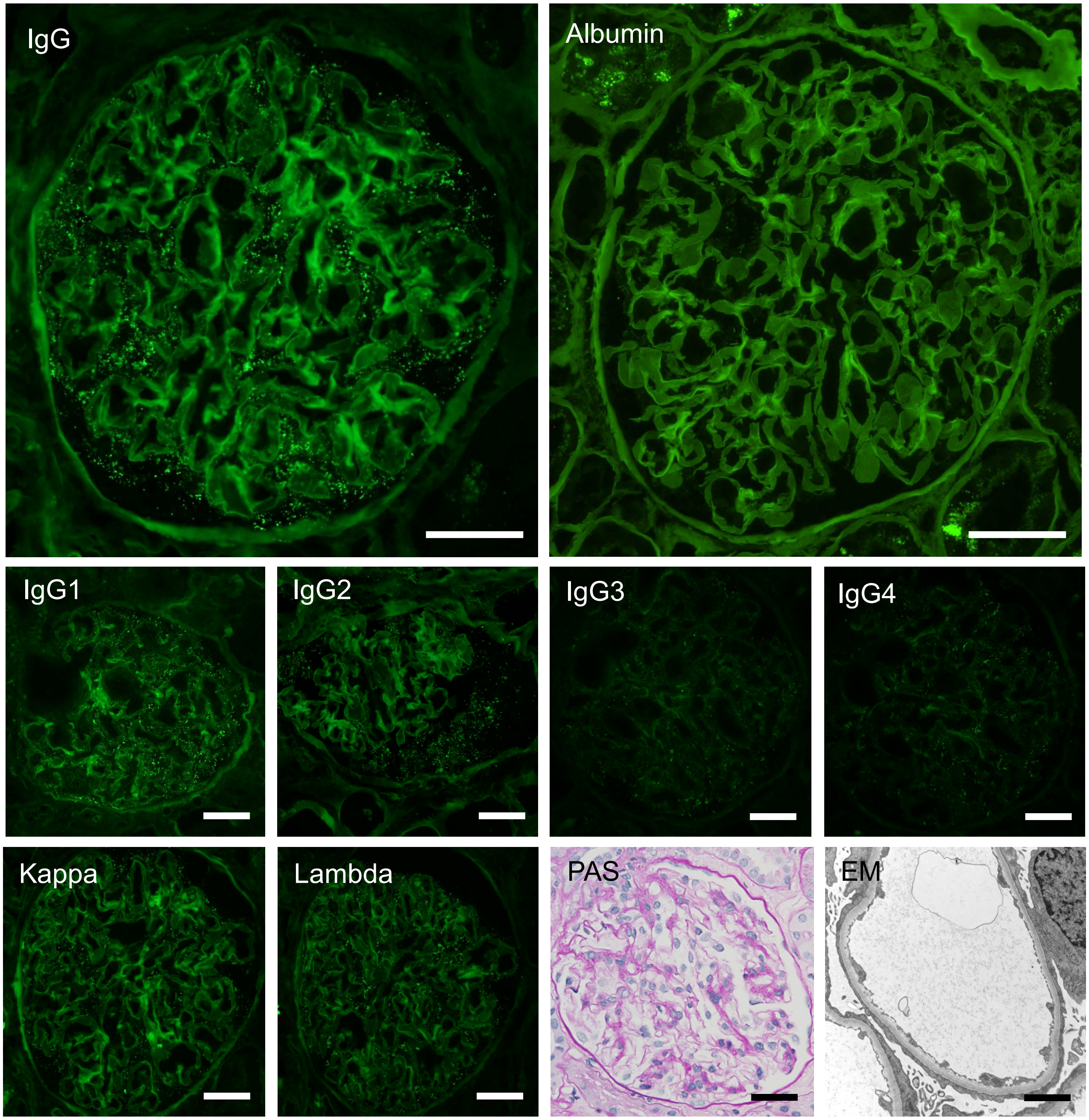


**Figure S6. Routine clinical epifluorescence microscopy, Periodic Acid Schiff (PAS) stained light microscopy and electron microscopy (EM) images of IgG-positive MCD (MCD7+).** The diffuse punctate staining seen with FITC-conjugated IgG antibody is not seen by albumin staining. Note the positive albumin staining in proximal tubule reabsorption droplets. Staining for IgG subtypes confirms no restriction of the punctate staining, and, in this case, more immunoreactivity for IgG1 and IgG2 compared to IgG3 and IgG4 subtypes was observed. Staining for Kappa and Lambda light chains shows equal intensity and an appearance mirroring the IgG staining. PAS staining shows minimal light microscopic changes and EM demonstrates diffuse podocyte foot process effacement. Scale bars IF and PAS: 20$\mu$m. Scale bar EM: 1$\mu$m.

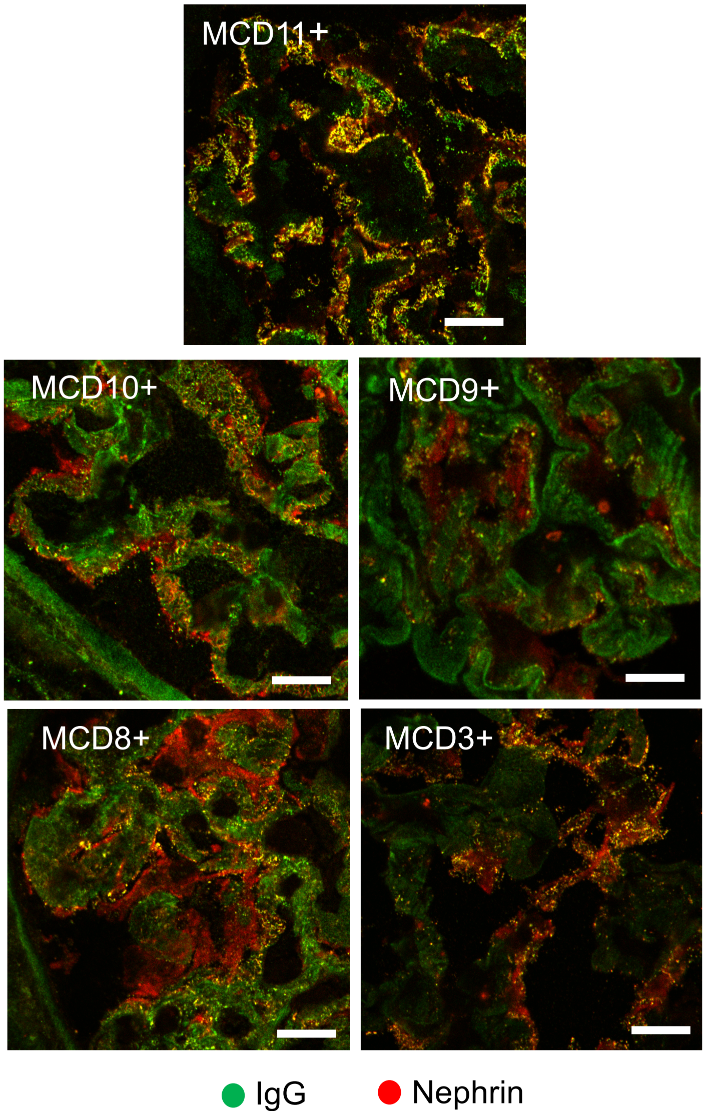


**Figure S7. Representative confocal microscopy images of 5 additional IgG-positive MCD renal biopsies showing colocalization between IgG and nephrin.** Immunofluorescence staining showing colocalization (yellow) between IgG (green) and nephrin (red) in renal biopsies from patients presenting with acute onset nephrotic syndrome (NS) and a histological diagnosis of minimal change disease (MCD) with diffuse foot process effacement and absence of electron-dense deposits by electron microscopy. Scale bar: 10µm

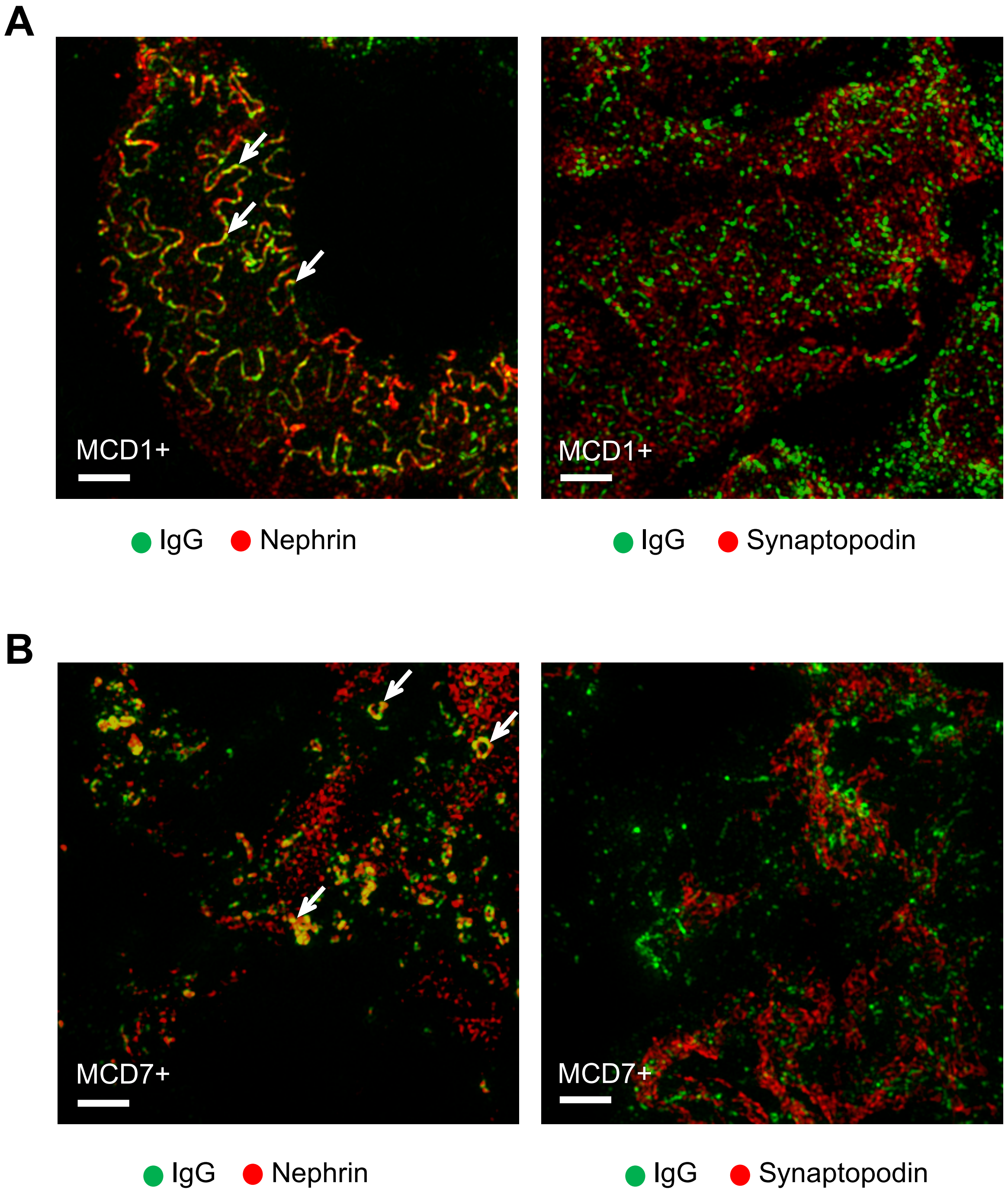


**Figure S8. Structured Illumination Microscopy images of representative MCD+ biopsies illustrating the two different patterns of IgG distribution observed in all MCD+ cases.** Complete Z-stack of multiple images taken at different focal distances (z-stack) combined to show greater depth of field for the biopsies shown in Figure 2 c, d. The two cases shown are representative of the two different patterns of IgG distribution observed in all MCD+ cases. (A) MCD1+ illustrates the co-localization (yellow) of IgG (green) with nephrin (red) in a pattern reflecting their presence along cell-cell junction (white arrows) (left). Essentially no overlap of the IgG (green) with the actin binding protein synaptopodin (red) is observed (right). (B) MCD7+ shows overlap (yellow) of IgG (green) with nephrin (red) in a more scattered granular, vaguely vesicular pattern (white arrows) (left). There is no overlap with the actin-binding protein synaptopodin (right). Scale bar: 2 μm

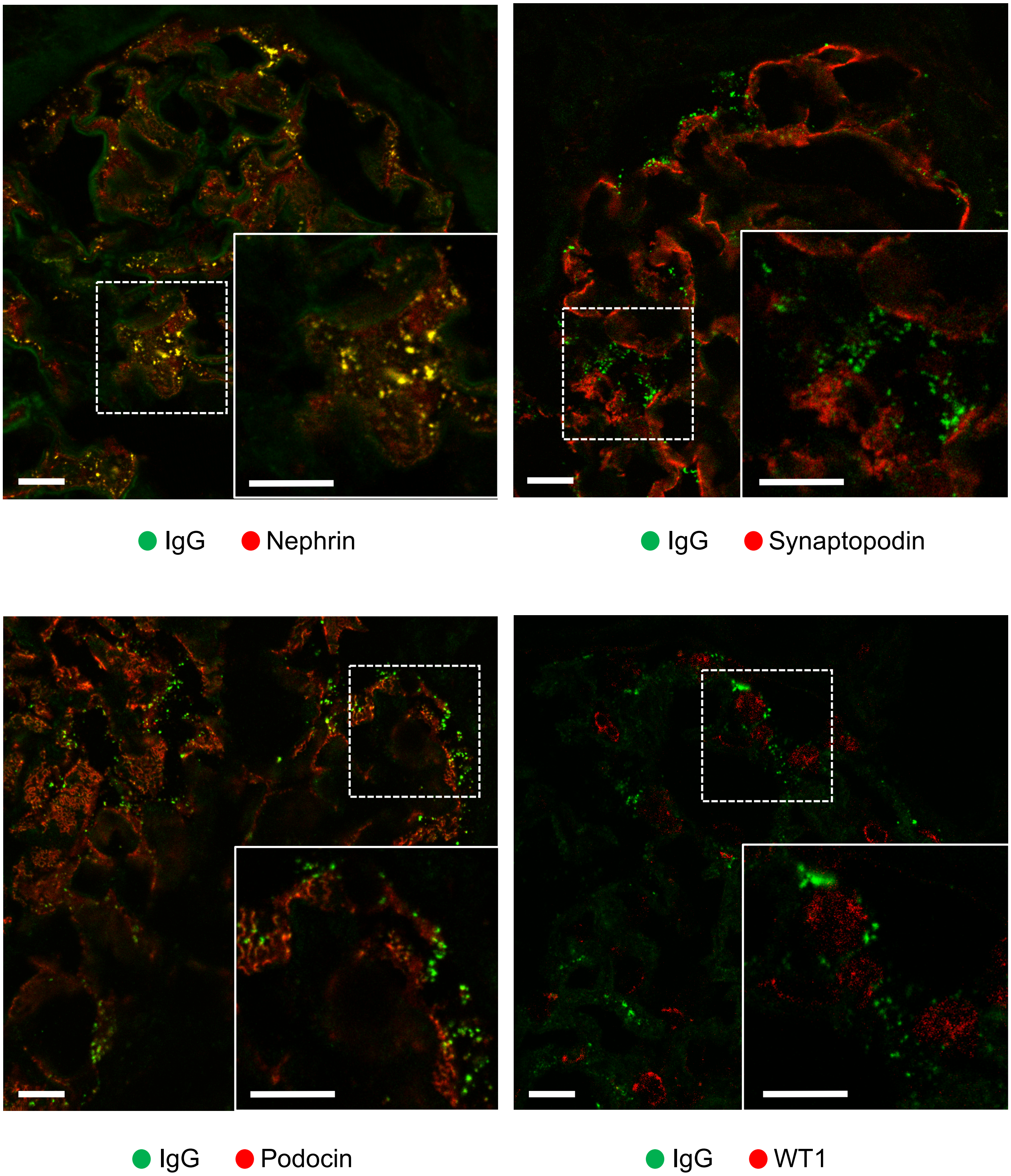


**Figure S9. Consistent co-localization of granular IgG with nephrin but not with other podocyte specific proteins in IgG-positive MCD.** Confocal microscopy imaging of a representative MCD+ biopsy (MCD12+) with granular IgG staining (green) illustrating substantive overlap (yellow) of IgG (green) with nephrin (red), but not with the cytoplasmic actin-associated foot process component synaptopodin (red), the intracellular slit diaphragm protein podocin (red) or the nuclear transcription factor WT1 (red). A magnified image of the boxed area (dotted line) is shown at the bottom right of each image (box with solid line). Scale bars: 5 µm

***BWH/MGH/BMC/Mayo Clinic cohort***

| Patient | Diagnosis | Age (5yr range) | Sex | Race/ Ethnicity | Proteinuria | Serum Cr (mg/dL) | $\alpha$–Nephrin Ab (U/ml) | Punctate IgG on biopsy |
|---|---|---|---|---|---|---|---|---|
| MCD1 (+) | Minimal change disease | 60 | Female | White | 28 | 2.2 | No serum | Yes (IgG1) |
| MCD2 (+) | Minimal change disease | 25 | Male | White * | 22.4 | 1.93 | 705 ± 103 | Yes (IgG1) |
| MCD3 (+) | Minimal change disease | 65 | Male | White | 7.32 | 2.81 | 5450 ± 1558 | Yes (IgG4) |
| MCD4 (+) | Minimal change disease | 30 | Male | Asian | 20 | 1.95 | 1375 ± 253 | Yes (IgG1) |
| MCD5 (-) | Minimal change disease | 70 | Female | White | 4.95 | 0.99 | No serum | No |
| MCD6 (-) | Minimal change disease | 50 | Male | White | 2.7 | 4.4 | 10 ± 12 | No |
| MCD7 (+) | Minimal change disease | 60 | Female | White | 12.95 | 1.4 | 330 ± 18 | Yes (IgG1) |
| MCD8 (+) | Minimal change disease | 60 | Male | White | 10.21 | 2.3 | 1427 ± 145 | Yes (IgG4) |
| MCD9 (+) | Minimal change disease | 40 | Female | Black | 18 | 2.45 | No serum | Yes (IgG1) |
| MCD10 (+) | Minimal change disease | 50 | Male | White | 8.33 | 1.2 | No serum | Yes (ND) |
| MCD11 (+) | Minimal change disease | 10 | Female | White | 3+ | 0.5 | No serum | Yes (IgG4) |
| MCD12 (+) | Minimal change disease | 30 | Male | Unknown | 5.8 | 0.95 | No serum | Yes (Ig4) |
| MCD13 (+) | Minimal change disease | 50 | Male | White | 15 | 2.4 | 649 ± 30 | Yes (ND) |
| MCD14 (+) | Minimal change disease | 20 | Female | White | 8.71 | 1.3 | 270 ± 50 | Yes (IgG4) |
| MCD15 (+) | Minimal change disease | 70 | Male | White | 7.2 | 2.55 | 911 ± 83 | Yes (IgG4) |
| MCD16 (+) | Minimal change disease | 80 | Male | White | 2 | 1.3 | 271 ± 39 | Yes (ND) |
| MCD17 (-) | Minimal change disease | 35 | Female | White | 2[#] | 0.95 | 36 ± 11 | No |
| MCD18 (-) | Minimal change disease | 80 | Male | White | 2.14 | 3.22 | 21 ± 3 | No |
| FSGS1- | Primary FSGS | 40 | Female | White | 4.19 | 0.51 | 0 ± 0 | No |
| FSGS2- | Primary FSGS | 55 | Male | White | 8 | 3.5 | 8 ± 6 | No |
| TL- | Podocytopathy with TL | 40 | Male | White | 10 | 1.02 | 94 ± 100 | No |
| Amyloid | Amyloidosis | 45 | Male | White | 12 | 3.01 | 7 ± 7 | No |
| MN1 | $PLA_2R$+ MN | 70 | Male | White | 7.6 | 0.87 | 24 ± 33 | ND |
| MN2 | $PLA_2R$- MN | 40 | Male | White | 0.83 | 0.87 | 16 ± 15 | ND |
| DN1 | Diabetic Nephropathy | 70 | Male | Unknown | 3.5 | 0.8 | No serum | No |
| DN2 | Diabetic Nephropathy | 40 | Male | White | 7.41 | 4.6 | 0 ± 0 | No |
| DN3 | Diabetic Nephropathy | 50 | Male | Black | 10 | 2.8 | 0 ± 0 | No |
| Nx1 | Nephrectomy for RCC | 80 | Male | White | 0.07 | 1.46 | 47 ± 32 | No |
| Nx2 | Nephrectomy for RCC | 60 | Male | White | negative | 1.51 | 19 ± 33 | No |
| Normal | Normal | 25 | Female | White | 0.2 | 0.74 | 32 ± 37 | No |

**Table S3. BWH/MGH/BMC/Mayo Clinic cohort clinical characteristics.** The BWH/MGH/BMC/Mayo Clinic cohort consists of patients whose renal biopsy was evaluated for IgG by immunofluorescence staining (IF) and a concurrent serum sample, where available, was evaluated for anti-nephrin antibodies. For the control patients that had a tumor nephrectomy for RCC (Nx1, Nx2) an area of non-neoplastic renal parenchyma was evaluated by IF. Proteinuria values are given as either UPCR (g/g) or urine dipstick (negative, 3+, 4+) unless otherwise stated ([#]For patient MCD17-, proteinuria is given as urine albumin creatinine ratio (UACR) (g/g)). Serum Creatinine (Serum Cr) and proteinuria values are those closest to the time of serum sampling for patients evaluated for anti-nephrin antibodies and closest to the biopsy for those who were not. The predominant IgG subclass is given in parenthesis where known (ND indicates that the IgG subclass was not determined due to lack of additional biopsy material). FSGS, focal segmental glomerulosclerosis; TL, tip lesion; MN, membranous nephropathy; RCC, renal cell cancer. * indicates Hispanic or Latino ethnicity.

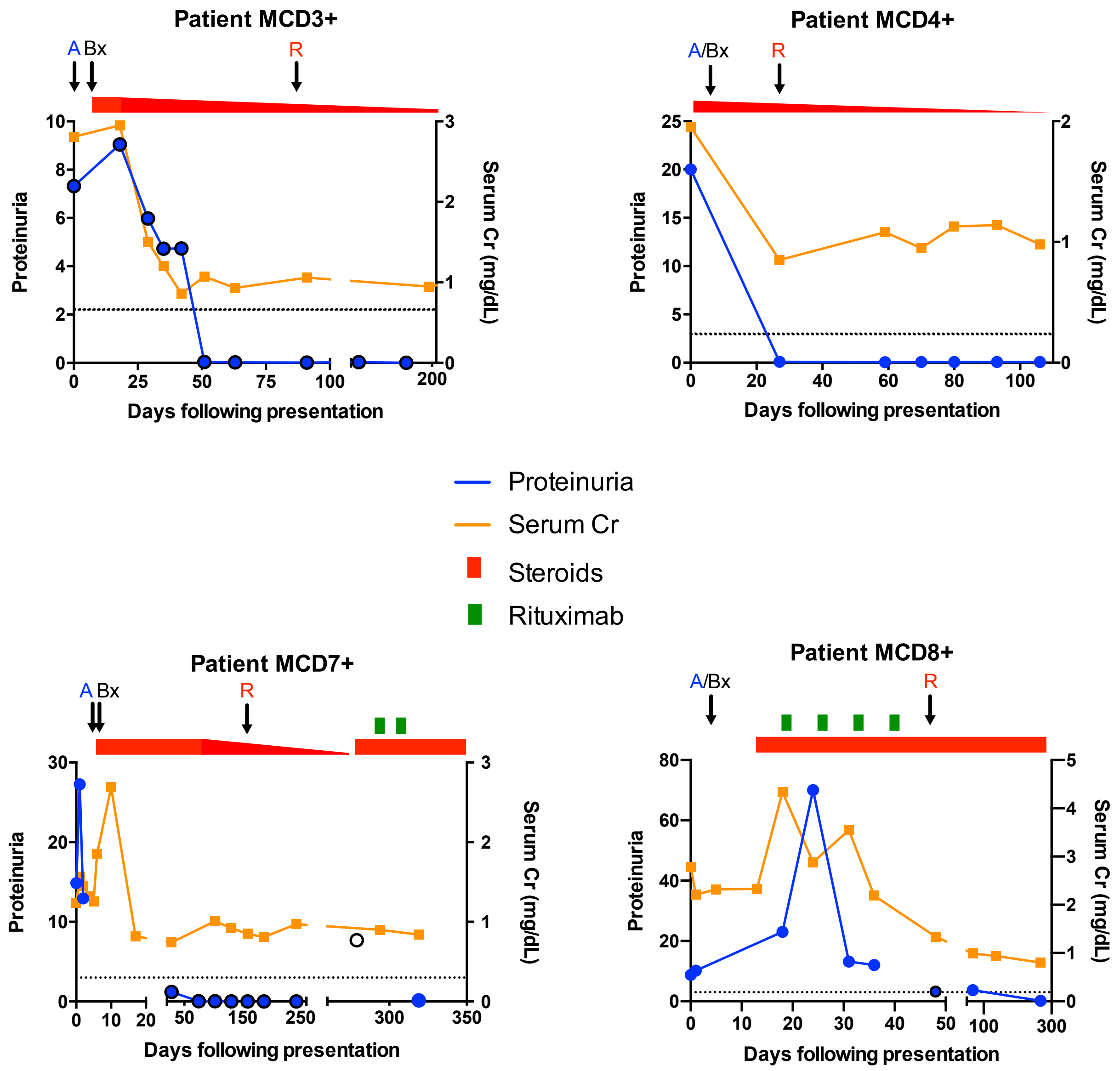


**Figure S10. Clinical course of biopsy IgG-positive MCD patients evaluated for nephrin autoantibodies during active disease and following response to treatment**. Proteinuria (Urine protein creatinine ratio, UPCR (g/g), blue circle; Urine albumin creatinine ratio, UACR (g/g), blue circle with black border; 24hr urine protein (g/24hr), clear circle with black border) and serum creatinine (Cr) are shown for patients with biopsy IgG-positive MCD (MCD+) whose serum was evaluated for anti-nephrin antibodies during active disease and during treatment response. Active samples (A) were obtained within 7-days of clinical presentation and response samples (R) were obtained when the patients were in complete (MCD4+, MCD7+) or partial remission (MCD8+), defined by UPCR < 0.3 g/g (UACR < 0.2 g/g) or 50% reduction in proteinuria respectively, on the day of sample collection. For MCD3+, the responsive sample was collected approximately 3 weeks after entering a period of sustained clinical remission. Treatment is shown at the top of the graph, with a dose reduction in steroids indicated by a downward sloping wedge. The dotted line indicates a proteinuria level of 3. Bx, renal biopsy.

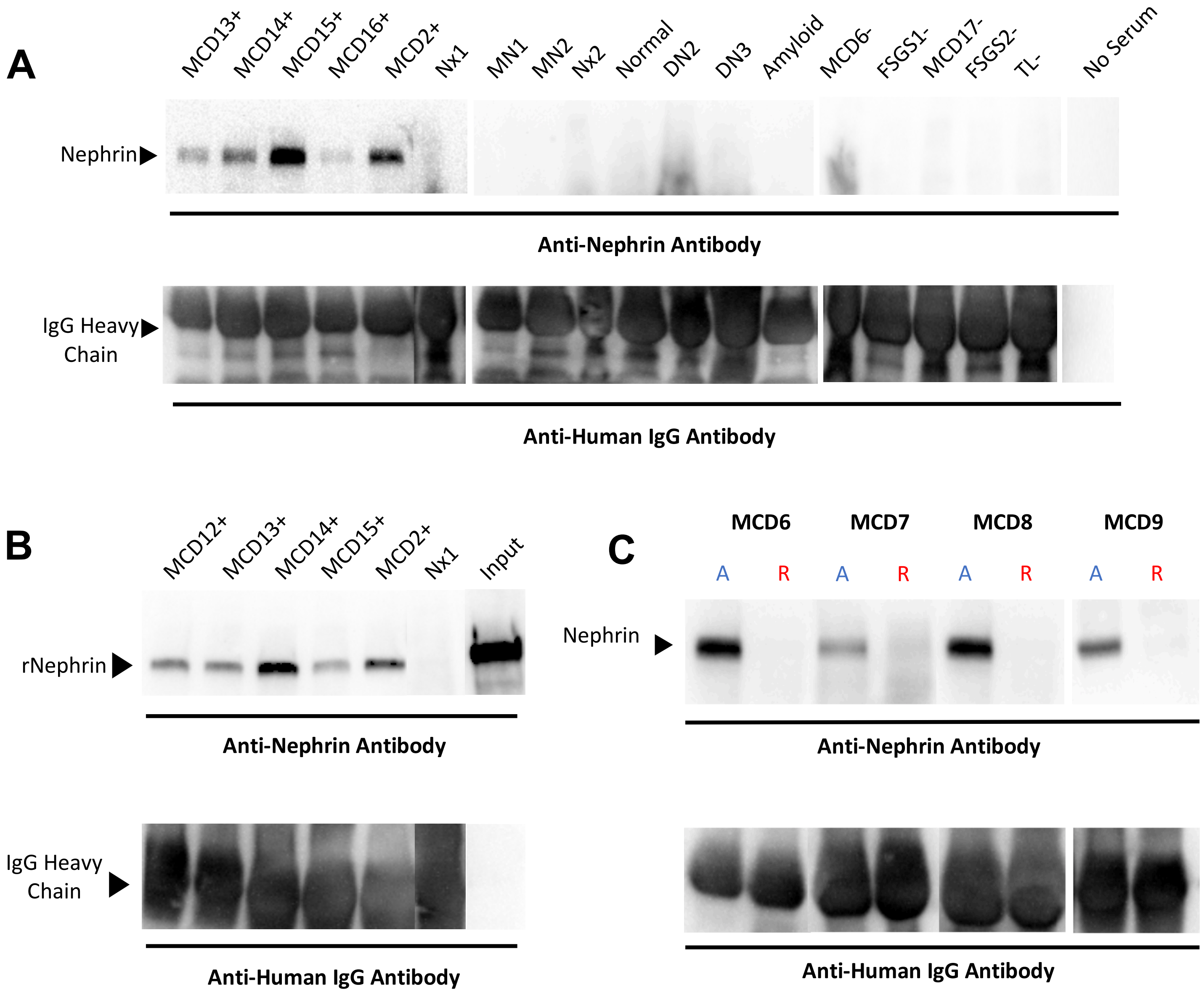


**Figure S11. Biopsy IgG+ MCD patient serum or plasma immunoprecipitates both nephrin from human glomerular extract (HGE) and affinity-purified recombinant extracellular domain of human nephrin** (A) Nephrin was immunoprecipitated from human glomerular extract (HGE), derived from healthy donor kidney, with serum from patients with biopdy IgG positive MCD (+) and not from control patients lacking IgG on renal biopsy. (B) Purified recombinant extracellular domain of human nephrin (hNephrin$_{G1059}$) was immunoprecipitated by serum or plasma from patients with MCD and punctate IgG in their renal biopsies (MCD2+, 13+, 14+, 15+, 16+), but not by a control patient without IgG deposition in the biopsy (Nx1, disease-free area of tumor nephrectomy). The input lane shows the starting amount of recombinant hNephrin$_{G1059}$ protein used for the immunoprecipitation (not incubated with serum or Protein G beads). C) Nephrin was precipitated from HGE in four MCD+ patients during active disease, but not following remission. Immunoprecipitates were electrophoresed under reducing conditions and subjected to Western blot analysis with a primary sheep anti-human nephrin antibody and secondary HRP-conjugated donkey anti-sheep IgG antibody (top) or a primary HRP-conjugated donkey anti-human IgG alone (bottom).

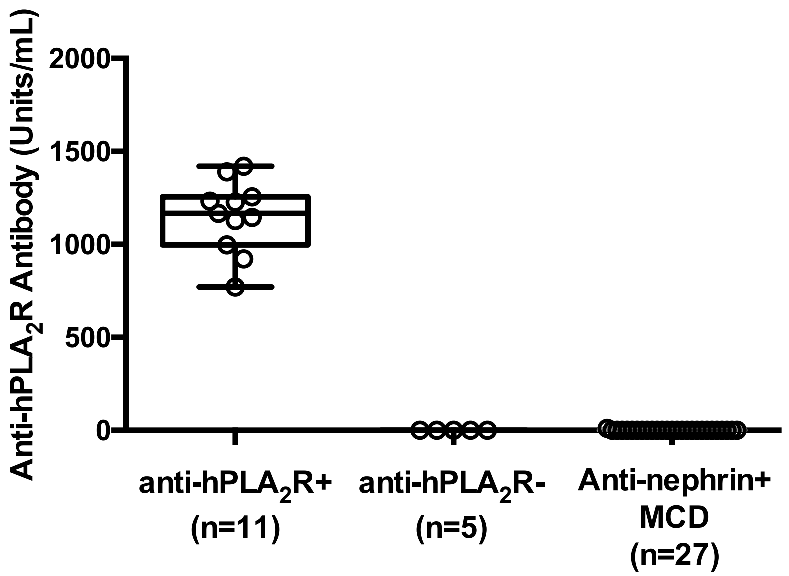


**Figure S12. Antibodies to nephrin in MCD patients do not cross react with $hPLA_2R$.** Serum from patients with anti-nephrin antibodies were evaluated for antibodies to the extracellular domain of recombinant affinity purified human $PLA_2R$ ($hPLA_2R$) by ELISA. Control serum was obtained from patients screened for anti-$hPLA_2R$ antibodies by two validated routine clinical assays; ELISA and IIFT (Euroimmun). The standard curve was generated using a single high titer patient sample with a 1:40,000 dilution defined to contain 1000 Units/ml.

**CLINICAL CASE DETAILS (pertaining to Figure S13)**

Per Medrxiv guidelines, this section was removed from the manuscript. Please contact the corresponding author for more information.

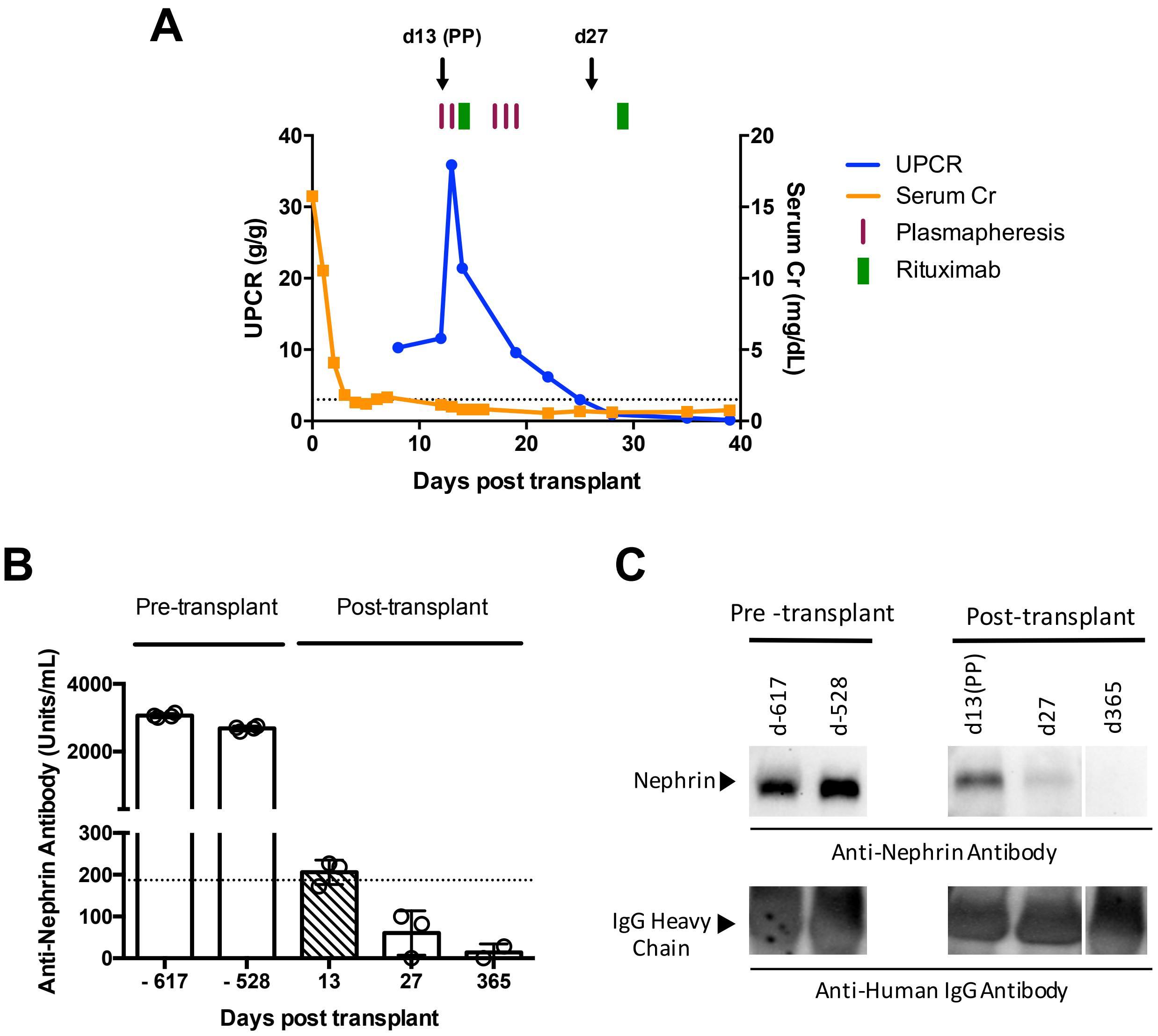


**Figure S13. High levels of pre-transplant anti-nephrin antibodies are associated with early massive post-transplant proteinuria recurrence**. (A) Clinical course of a patient with childhood onset, steroid dependent MCD that progressed to ESKD with subsequent biopsies showing FSGS. The patient developed early, massive proteinuria recurrence, following a cadaveric, pediatric donor kidney transplant that rapidly responded to treatment with plasmapheresis (purple lines) and rituximab (green lines) shown above the graph. (B) Two pre-transplant serum samples (day -617, day -528) and the initial plasmapheresate (shaded box) (d13 (PP), indicated by arrow on (A)) tested positive by ELISA for anti-nephrin antibodies (dotted line indicates threshold for positivity of 187 U/ml). Serum samples evaluated following treatment response at d27 (d27, indicated by arrow in (A)) and d365 (UCPR < 0.3 g/g) were negative for anti-nephrin antibodies. (C) Similarly, the pre-transplant serum and plasmapheresate immunoprecipitated nephrin from HGE (derived from disease free human kidneys) in keeping with the ELISA findings.
